# Occupational Pesticide Exposure and Regional Brain Volume Differences in UK BioBank

**DOI:** 10.64898/2025.12.22.25342092

**Authors:** Shane Johnson, Adam Kehl, Jonathen Ahern, Mary E.T. Boyle, Robert Loughnan

## Abstract

**Background:** Parkinson’s Disease (PD) is a progressive neurodegenerative disorder influenced by both genetic and environmental factors. Prolonged exposure to pesticides has been proposed as an environmental risk factor, yet its relationship with structural brain differences in human adults remains unexplored.

**Objective:** To investigate the association between occupational pesticide exposure and volumetric differences in specific brain regions, and to evaluate the relationship between pesticide exposure and PD diagnosis using data from UK BioBank.

**Methods:** Structural MRI (T1-weighted scans) were used to extract regional brain volumes. The study population contained 21,049 UK BioBank participants, of which 912 participants indicated they had occupational pesticide exposure. General linear models were utilized to assess the association between pesticide exposure and regional brain volume differences, adjusting for participant demographics, socioeconomic status, study site where imaging was conducted, and genetic principal components as covariates. A Cox proportional hazards model was utilized to assess the association between pesticide exposure and PD diagnosis (ICD-10 code G20).

**Results:** Significant negative associations were observed between occupational pesticide exposure and volumes of total gray matter, cortex, thalamus, ventral diencephalon, cerebellum cortex, cerebellum white matter, and brainstem. Significant positive associations were observed between occupational pesticide exposure and white matter hyperintensities and inferior lateral ventricle volumes. No significant hemispheric lateralization was observed, and adjusting for fluid intelligence scores did not significantly change model estimate values, demonstrating brain volumetric and pesticide associations were not confounded by differences in fluid intelligence. A trending, non-significant association between pesticide exposure and PD diagnosis was observed (Odds Ratio (95% CI) 1.36 (0.98 to 1.90) p=<0.12).

**Conclusion:** Occupational pesticide exposure is associated with structural reductions in volume within several brain regions, and increases in white matter hyperintensities and ventricular volume, supporting a possible neurodegenerative effect. This is the first large scale study to link adult pesticide exposure to region-specific brain volume differences utilizing non-invasive structural neuroimaging. These findings highlight a putative link between pesticide exposure and gross neuroanatomical changes in adulthood which may impact overall brain health.

## Introduction

Parkinson’s Disease (PD) is one of the most prevalent and fastest growing neurodegenerative disorders, characterized by motor symptoms that include tremor, rigidity, dystonia, slowness of movement, and postural instability (1,2). Research surrounding PD supports a complex relationship between an individual’s genetic predispositions and environmental factors that dictate the likelihood for onset of PD itself, or clinical symptoms that manifest themselves under the clinical syndrome of Parkinsonism (2,3,4). PD has long been characterized as selective loss of dopaminergic neurons in the substantia nigra, which projects to the basal ganglia (referred to as the nigrostriatal system), with autopsy findings suggesting that alpha-synucleinopathies and tauopathy proteins accumulating in degenerating neurons and glia are among the most common neurodegenerative causes of Parkinsonism. Other notable brain regions heavily involved in PD and Parkinsonism include the cerebellum, supplementary motor complex, prefrontal cortex, and parietal and premotor cortices (3, 4, 5, 6).

Occupational pesticide, insecticide, fungicide, and herbicide exposure has been linked to various diseases, among which include PD and the clinical syndrome, Parkinsonism (7, 8, 9, 10). These environmental contaminants have been shown to disrupt normal cell functions through a variety of mechanisms (i.e. inhibiting acetylcholinesterase, disrupting mitochondrial respiration, and introducing oxidative stress), and certain herbicides, such as Paraquat and Rotenone, have been strongly associated with PD diagnosis due their selective targeting of dopaminergic neurons in the nigrostriatal system (11, 12). In fact, agricultural records show that long-term exposure to a subset of pesticides are associated with PD, with several of these pesticides showing direct toxicity to regionally selective dopaminergic neurons (12, 13). Another geographical analysis displays a strong correlation between early age environmental pesticide exposure and the development of idiopathic Parkinson’s disease (14). As such, various neurotoxic chemicals and factors related to pesticide exposure have been proposed as potential risk factors for progressive symptoms related to Parkinsonism and PD.

Existing literature surrounding the possible relationship between pesticide exposure and volumetric differences in brain structure highlights conflicting results. Organophosphate (OP) concentration measured during pregnancy was found to have no statistically significant effect on cortical thickness, surface area, and brain volume, but was found to be associated with decreased white matter microstructure integrity in preadolescents (15). In contrast, one study in adults has shown that exposure to OPs during warfare, was associated with reductions in total gray matter and hippocampal volume (16), suggesting that the cortical effects of pesticides may differ depending on the developmental timing and dosage of exposure. A related study in tobacco farmworkers found no significant volumetric differences, as assessed using modulated voxel based morphometry, related to occupational pesticide exposure (17). These conflicting findings in adults may be due to limited sample sizes or degree/toxicity of exposure - e.g. warfare vs occupational exposure. Functional studies in children have revealed behavioral and executive function activation are alterations in those with prenatal OP exposure (18, 19, 20). Furthermore in animal models, OP exposure is linked with decreased brain volume in the frontal regions, decreased white matter integrity, and significant spatial learning deficits (21). In the current study we aim to resolve some of the conflicting findings in adults by conducting a structural brain imaging analysis to assess the effect of pesticide exposure in a sample over 100 times larger than these previous studies. We additionally aim to further assess the association between pesticide exposure and PD diagnosis within adult human participants. Given previous findings we hypothesized that, in adults, occupational pesticide exposure would either be related to: a) gross anatomical reductions in total cortical gray matter and hippocampal volume - as found in veterans exposed to OP (16), or b) no volumetric differences as observed previously in farmworkers (17). Furthermore, we anticipated that occupational pesticide exposure would be associated with increased Parkinson’s disease risk as demonstrated by a wealth of previous literature.

## Methods

### UK BioBank Sample

UK BioBank is a population-based cohort consisting of 502,150 participants aged between 40-69 years, with extensive medical, lifestyle, and genetic data recorded for each individual (23). MRI scans, genotypes, demographic, diagnostic, and clinical data were obtained from the UK BioBank under accession number 27412. All participants provided electronic signed informed consent, and the study was approved by the UK BioBank Ethics and Governance Council. The recruitment period for participants was from 2006 to 2010. Health records, genotype, and neuroimaging data were collected from January 2006 to May 2024. Data analysis was conducted from April 2024 to May 2025 (22).

### Occupational Pesticide Exposure Questionnaire (UK BioBank)

Data for pesticide exposure responses were retrieved from UK BioBank data-field 22614. The occupational pesticide exposure questionnaire was administered on UK BioBank under the section titled ‘Work History’, in which for each profession, the subject was given the question: “Did you work with pesticides?” with the available responses being “Often”, “Sometimes”, “Do Not Know”, and “Never”. We coded the ‘Sometimes’ and ‘Often’ responses as a binary ‘1’ or ‘True’ value, and conversely coded the ‘Never’ responses as a binary ‘0’ or ‘False’ value, omitting individuals responding ‘Do Not Know’ from our analysis. Because this question was asked per job/profession throughout an individual’s career, we coded any instances of ‘Sometimes’ and ‘Often’ as a binary ‘1’ or ‘True’ value. Subjects did have the choice to skip this question. After the exclusion of participants that withdrew consent, there were a total of 119,811 individuals who answered the occupational pesticide exposure questionnaire, with a vast majority of participants indicating a ‘Never’ response (114,804 individuals) and 5,007 individuals indicating either a ‘Sometimes’ or ‘Often’ response.

### Image Acquisition

T1-weighted structural imaging provides information related to an individual’s brain tissue and structural volume and morphology, using contrasts between gray and white matter to reflect differences in the interaction of water with surrounding tissue (24). T1-weighted scans were collected from 4 scanning sites throughout the UK, all on identically configured Siemens Skyra 3T scanners, with 32-channel receiver head coils. T1 scans were collected using a 3-dimensional magnetization-prepared rapid gradient-echo sequence at 1-mm isotropic resolution.

Imaging-derived phenotypes from the T1-weighted scans and FreeSurfer (25, 26, 27, 28) were utilized to generate a score of the subcortical structure volume of brain regions of interest (ROI) from participants in UK BioBank. Field names, their corresponding names used within the figures, descriptions of each brain region and results of this analysis can be found in *Supplementary Table 1*. At the time of our analysis, 21,049 individuals had adequate T1-weighted imaging, and therefore ROI brain volume scores. There were 21,049 individuals that both answered the occupational pesticide exposure questionnaire and had ROI brain volume scores, with 912 individuals indicating a “True” value on the binarized occupational pesticide variable described above.

### Statistical Analysis

General linear regression models were fit to test the association between pesticide exposure and regional brain volume differences among T1-weighted scans. Each model was corrected for the following confounding variables: age at time of scan (data-field 21003), sex (data-field 31), top 10 principal components of genetic ancestry (data-field 22009), the clinical site the MRI scan was conducted (data-field 54), education score (data-field 26414), health score (data-field 26413), and income score (data-field 26411). We included the education, health, and income deprivation scores as confounding variables to account for socioeconomic status among participants. We also included the clinical site as a confounding variable to account for the different scanners used for imaging. The genetic covariates (C1-C10) represent ancestral differences that account for genetic variation among participants. Lastly, we also included estimated total intracranial volume (eTIV) so that effects could be interpreted as regional effects rather than effects resulting from differences in total brain volume.

Across all generalized linear regression models, we performed multiple comparisons corrections using both the Bonferroni method and Benjamini-Hochberg False Discovery Rate (FDR) method (29, 30). Nominal significance (p<0.05) among ROI volumes is indicated in the respective forest plots with a horizontal cyan blue line, significance after FDR correction is indicated with a horizontal green line, and significance after Bonferroni correction is indicated with the horizontal purple line. The z-scores for all ROI volumetric values were calculated before performing regressions, allowing coefficient values to be interpreted as normalized standard regression coefficients.

We calculated the average ROI volume across hemispheres to detect bilateral effects, which are labeled “ROI_avg” in the respective figures below. Additionally in supplementary analysis to detect lateralized effects, we took the difference between left and right hemispheres for each ROI and used these as the response variable of interest. Information regarding participant fluid intelligence scores as a confounding variable and ROI lateralization effects can be found in the Supplementary Methods section.

### Survival Analysis Using Parkinson’s Disease Diagnosis

We conducted a Cox proportional hazards model to assess the relationship between occupational pesticide exposure and PD diagnosis (event incidence). For the purposes of this survival analysis, a Parkinson’s disease diagnosis was characterized as ICD 10 code G20 (data-field 41202). We included sex, education scores, health scores, income scores, and top 10 principal components of genetic ancestry (C1-C10) as covariates. The total number of individuals who were diagnosed with Parkinson’s Disorder and answered the pesticide exposure questionnaire (as either ‘True’ or ‘False’) was 551 out of 101,957 individuals. There were 21,049 individuals that both answered the occupational pesticide exposure questionnaire and had ROI brain volume scores, with 912 individuals indicating a “True” value on the occupational pesticide questionnaire. For this survival analysis, the duration to the event (PD diagnosis) was calculated as either the date of birth to the date of diagnosis, or to the date of death/date of censor, where the date of death/censor do not reach PD diagnosis event. Further description of the UK BioBank sample used in this study can be found in *Supplementary Table 1*.

## Results

### Association between occupational pesticide exposure and volumetric differences across several ROIs

Figure 1 displays the association between occupational pesticide exposure and regional volumetric brain differences (including white matter hyperintensities) averaged across hemispheres to detect bilateral effects. Critically, we include total intracranial volume as a covariate in addition to study specific and sociodemographic covariates described in the Methods section. We observed significant negative associations, indicating smaller brain regions, between occupational pesticide exposure and the following ROIs (where coefficients are standardized regression coefficients): total gray volume (Coef (95% CI) -0.085, p=1.95e-07), average cortex volume (Coef (95% CI) -0.086, p=1.14e-06), average thalamus volume (Coef (95% CI) -0.068, p=2.31e-03), average ventral diencephalon volume (Coef (95% CI) -0.067, p=<2.67e-03), average cerebellum white matter volume (Coef (95% CI) -0.062, p=<2.24e-02), average cerebellum cortex (Coef (95% CI) -0.054, p=<3.62e-02), and brainstem (Coef (95% CI) -0.049, p=<4.5e-02). We also observed significant positive associations between occupational pesticide exposure and white matter hyperintensities (Coef (95% CI) 0.10, p=<1.43e-03), and average inferior lateral ventricle (Coef (95% CI) 0.056, p=<3.50e-02).

**Figure 1:**
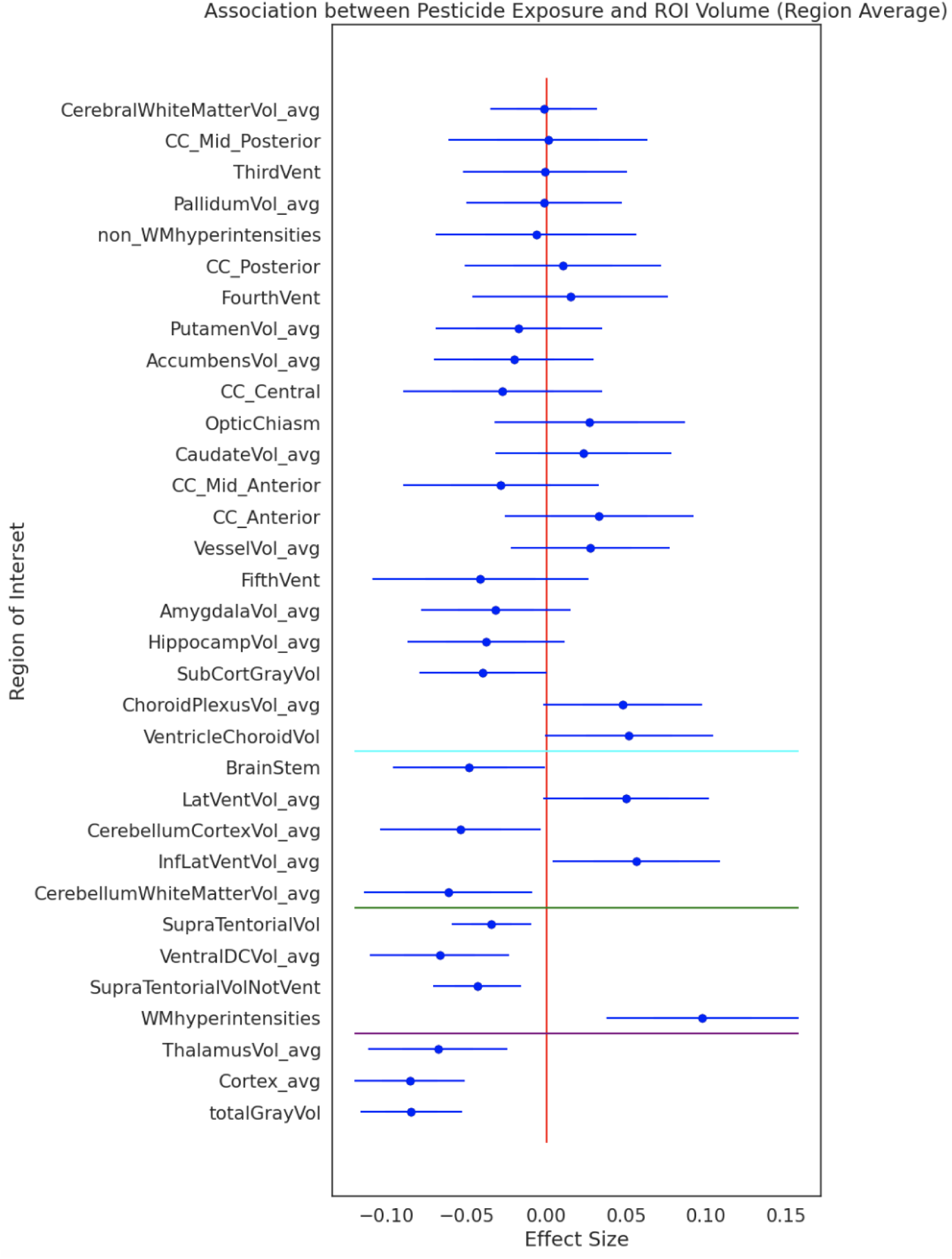
The association between occupational pesticide exposure and regional brain volume. Forest plot depicting the association between occupational pesticide exposure and average regional brain volumes (ROI) using generalized linear regression models. ROI volume values were averaged between hemispheres to account for bilateral effects. Standard covariates were utilized as mentioned in the Methods section, namely sex assigned at birth, age at the time of imaging, top 10 principal components of genetic ancestry (C1-C10), Indices of Multiple Deprivation (health, education, and income), clinic site where imaging was conducted, and estimated total intracranial volume. The horizontal blue line represents nominal statistical significance, the horizontal green line represents the significance threshold for False Discovery Rate (FDR) correction, and the horizontal purple line represents the significance threshold for Bonferroni correction. Coefficients and confidence intervals for all figures can be found below. An index for the ROI variables can be found in Table 2.

**Table 1:** Cohort Demographics (UK BioBank) Sample and participant demographics for both the imaging cohort, involved in imaging analyses involving ROI, and the non-imaging cohort, involved in the Cox proportional hazards model analysis.

|  |  |
| --- | --- |
| **Imaging Cohort - GLM w/ Multiple Corrections Analysis** |  |
| **Study Population** | 21,049 participants |
| **Average Age** | 64.49 years |
| **Male** | 9,820 participants |
| **Female** | 11,229 participants |
| **Total Number of PD Cases (G20)** | 51 diagnosed individuals |
| **Total Number of Responses to Pesticide Exposure Questionnaire** | 21,049 responses |
| **"Often" and "Sometimes" Responses (T)** | 912 responses |
| **"Rarely/Never" and "Do Not Know" Responses (F)** | 20,137 responses |
| **Non-Imaging Cohort - CPH Analysis** |  |
| **Study Population** | 101,957 participants |
| **Average Age (at Event)** | 56.68 years |
| **Male** | 45,357 participants |
| **Female** | 56,600 participants |
| **Total Number of PD Cases** | 551 diagnosed individuals |
| **Total Number of Responses to Pesticide Exposure Questionnaire** | 101,957 responses |
| **"Often" and "Sometimes" Responses (T)** | 4,250 responses |
| **"Rarely/Never" and "Do Not Know" Responses (F)** | 97,707 responses |

**Table 2:**
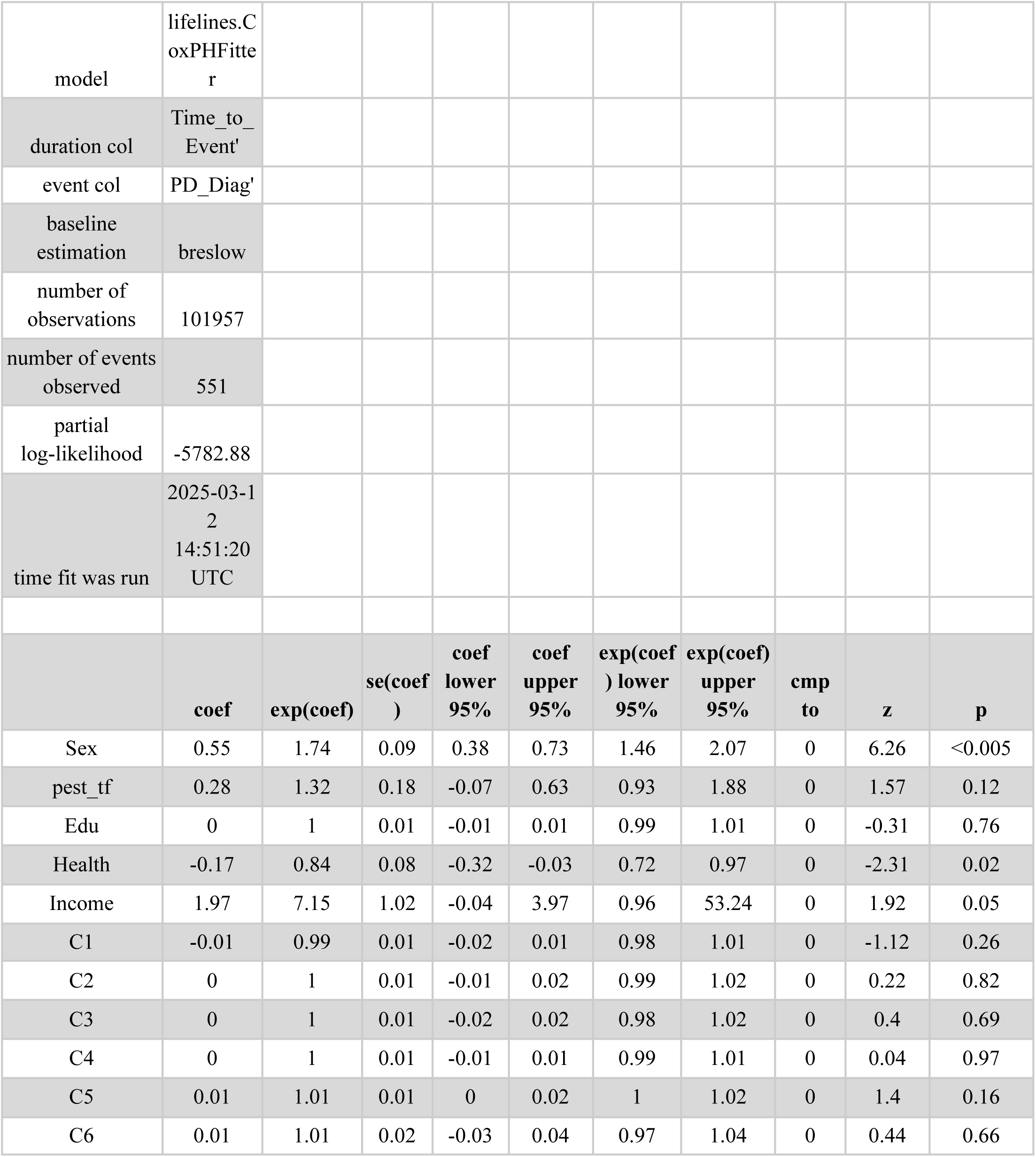
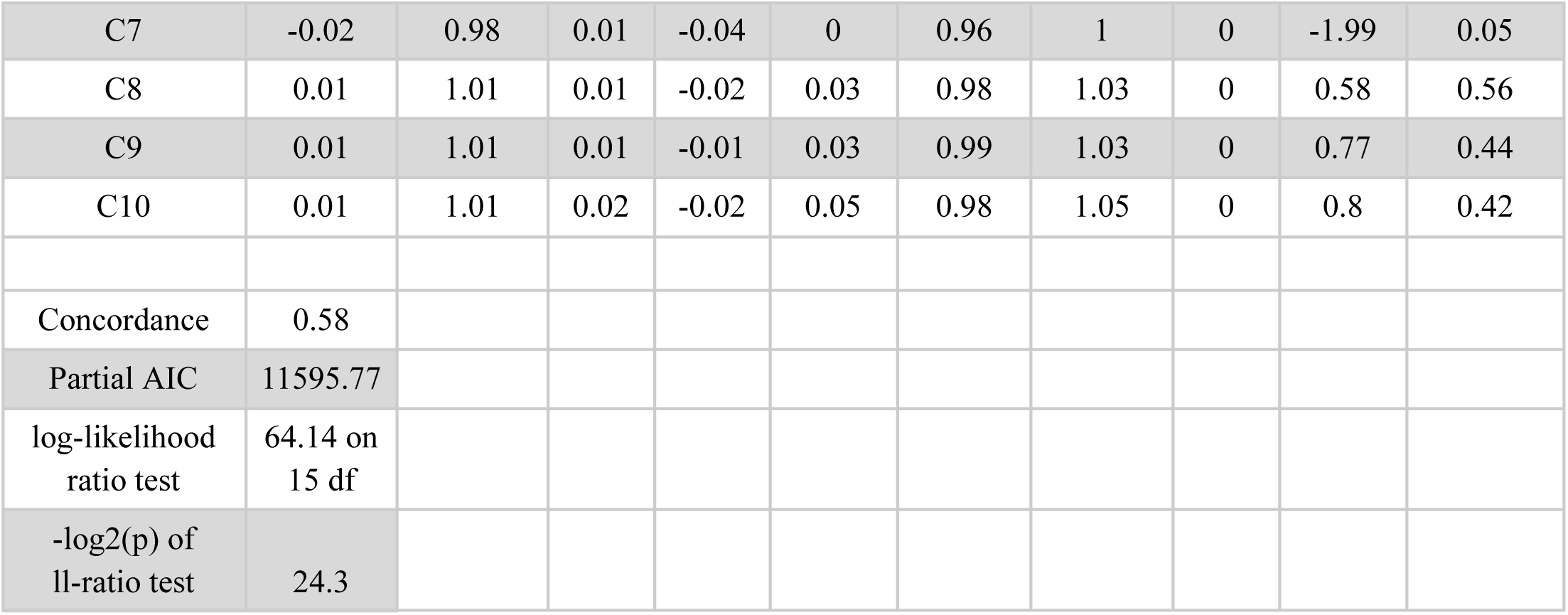
Using a Cox Proportional Hazard Model to analyze the association of pesticide exposure and PD diagnosis (G20) Model summary for a survival analysis using Cox proportional hazards model to test the association between occupational pesticide exposure and PD diagnosis (ICD 10 code G20). The duration to the event (PD diagnosis) was calculated as either the date of birth to the date of diagnosis, or to the date of death/date of censor, where the date of death/censor do not reach PD diagnosis event. We included sex, education scores (IMD), health scores (IMD), income scores (IMD), and top 10 principal components of genetic ancestry (C1-C10) as covariates in this analysis.

**Table 3:**
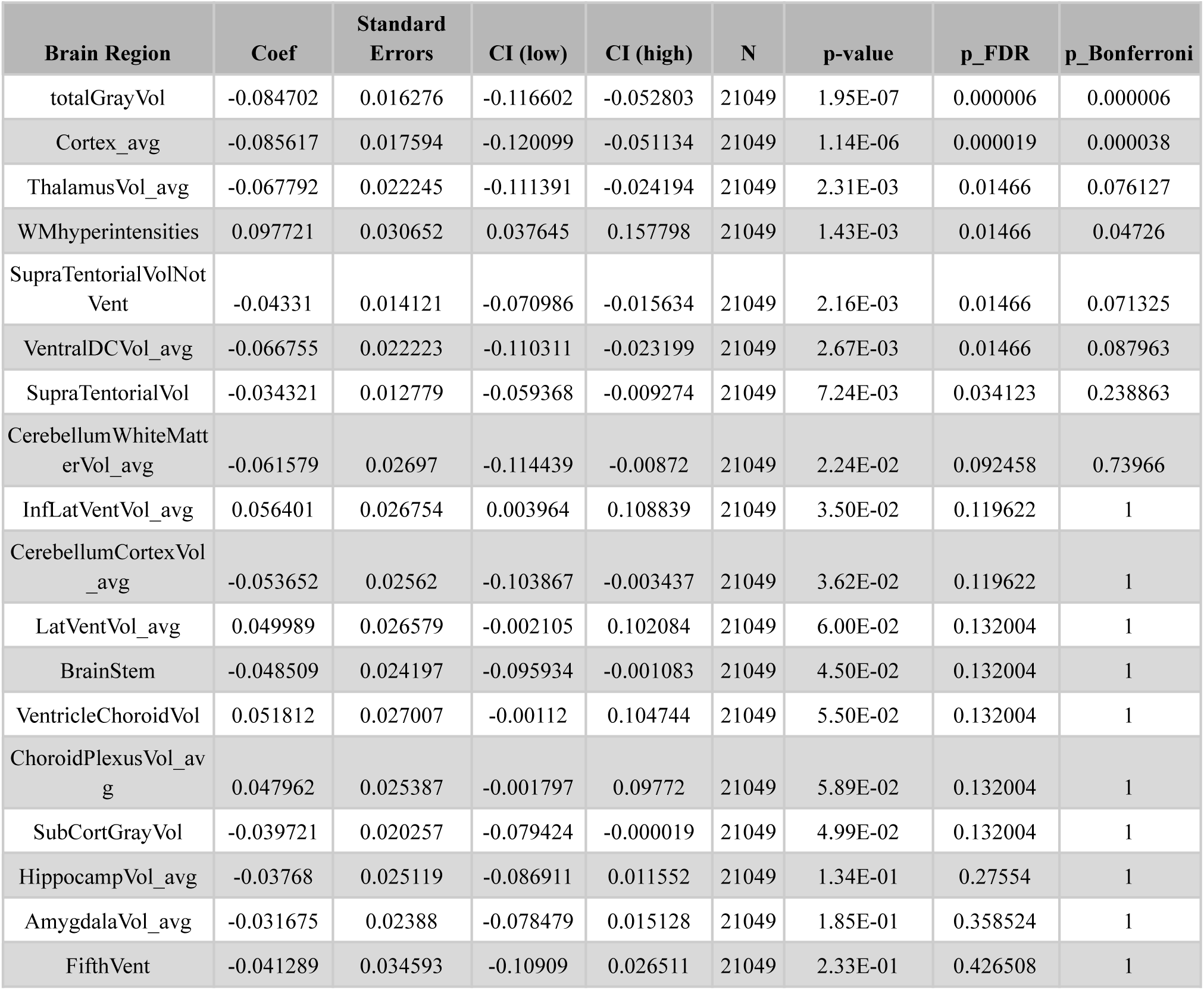
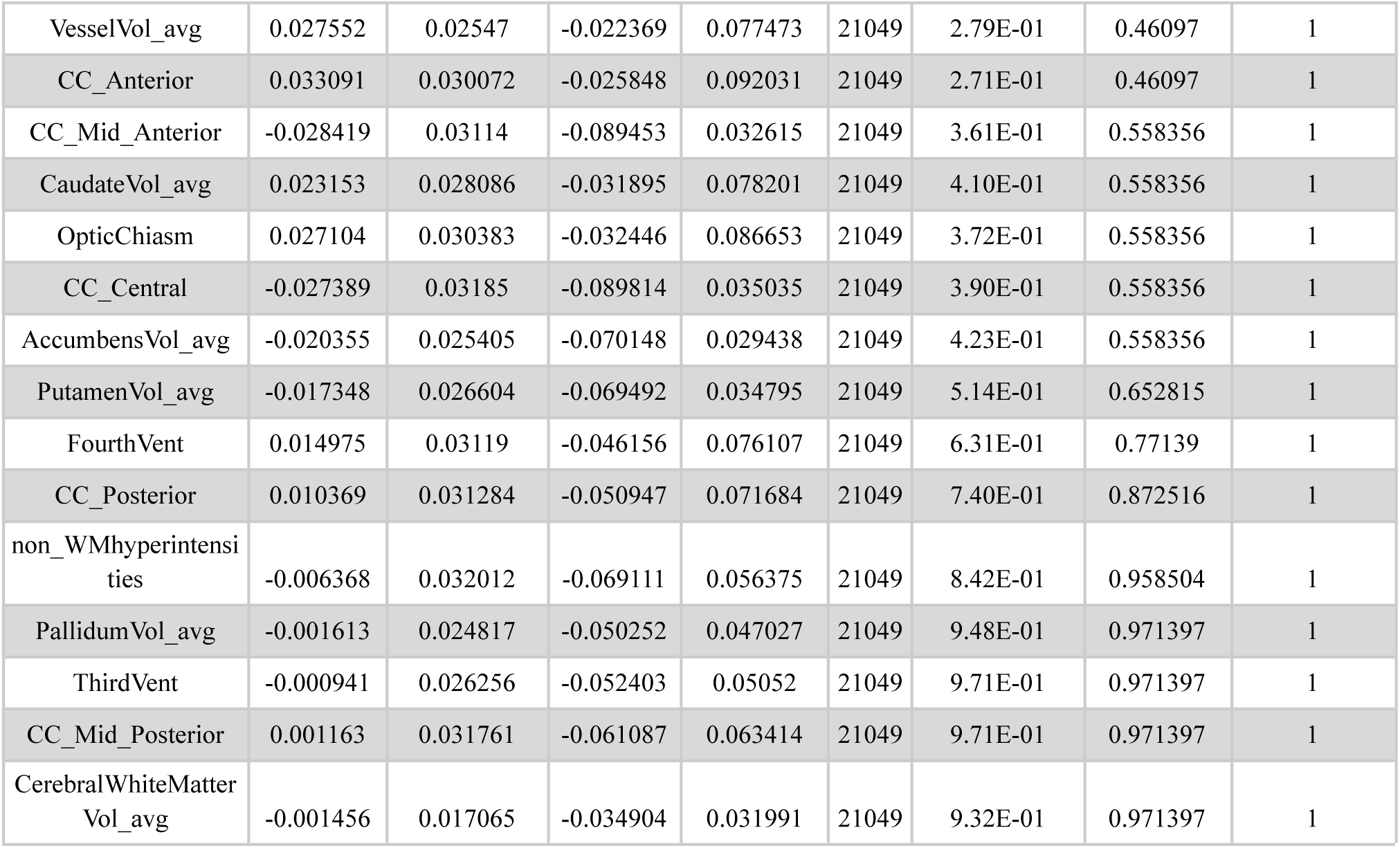
Corresponding Model Summary for Figure 1. Model summaries corresponding to the Figure 1 forest plot. Model summaries include the brain regions of interest (coded ROIs), model estimates (Coef), standard errors, low and high confidence intervals, the sample size (N), the nominal p-value, and the p-values of both FDR and Bonferroni multiple comparisons test for each ROI.

Supplementary Figure 1 displays model outputs from the analysis of volumetric differences between right and left hemisphere ROIs, for which we find no evidence of lateralized effects. In the Supplementary Analyses, we wanted to ensure that our significant global brain effects related to pesticide exposure were not being confounded with cognitive performance, therefore we performed regressions with the same covariates as Model 1, with the addition of fluid intelligence scores. The analysis revealed the model estimates remained largely unchanged (see Supplementary Figure 5 and 6) – indicating that cognitive performance differences did not have a large influence on the global brain effects we detected.

### The effect of occupational pesticide exposure on PD diagnosis using a Cox-proportional hazards model

Out of 101,957 individuals, there was a trending non-significant positive association between occupational pesticide exposure and PD diagnosis (Hazards Ratio (95% CI) 1.32, p=<0.12).

## Discussion

Our study of 21,049 participants revealed significant associations between occupational pesticide exposure and volumetric differences in specific brain regions, including white matter hyperintensities. No significant hemispheric lateralization effects were observed, suggesting that these significant associations are global, rather than hemisphere specific. Furthermore, adjusting for fluid intelligence scores did not substantially change model estimates, indicating that cognitive performance was not a confounding factor in the observed volumetric differences in global brain regions. Although our Cox proportional hazards model identified a trending positive association between pesticide exposure and PD diagnosis, this association did not reach statistical significance.

To the best of our knowledge, this is by far the largest study to investigate volumetric brain differences associated with occupational pesticide exposure. Two previously related studies in adults found conflicting results: with one study finding substantial reductions in total gray matter and hippocampal volume associated with exposure to OP in warfare (16), and another finding no volumetric differences associated with occupational pesticide exposure for farmworkers (17) - as measured by modulated voxel based morphology. Although we did not observe reductions in hippocampal volume, our findings align more closely with prior reports in veterans showing reduced total cortical gray matter. Notably, the effect size in our study was about 60% smaller, which could reflect differences in either the dosage or the specific toxicities of OP exposures experienced by veterans versus farmworkers. Such differences in exposure intensity or chemical profile may also account for the absence of hippocampal effects in our sample. Alternatively, exposure to chemical agents in warfare may have served as a proxy for severe warfare conditions which could have additionally resulted in brain volume reductions. We suspect that the null finding in the previous agricultural worker study may be a combined result of reduced statistical power compared with our study and a reduced toxicity and/or dosage of OP exposure when compared with the veterans study. Taken together we find these results underscore the plausibility that acetylcholinesterase-inhibiting pesticides can affect brain structure morphology.

As mentioned previously, pesticides, herbicides, and insecticides are known to disrupt neuronal integrity through several mechanisms, including inhibition of acetylcholinesterase, mitochondrial dysfunction, induction of oxidative stress, and promotion of neuroinflammation (12, 20, 31, 32, 33, 34, 35, 36). The precise mechanism varies depending on the chemical composition of the environmental pollutant, which may explain regionally selective neurotoxic effects observed in experimental models (37). Paraquat, for example, crosses the blood-brain barrier via DAT transporters and selectively targets dopaminergic neurons in the substantia nigra, a hallmark region in PD (12, 38). A recent pesticide-wide association study identified approximately ten pesticides capable of inducing substantial dopaminergic neuron death in the midbrain at specific concentration thresholds, further supporting the biological plausibility of our findings (13). Despite prior reports, we did not detect any effects in nigrostriatal regions. This does not rule out the possibility that OP exposure causes toxic damage there; rather, if such damage exists, it may not manifest as volumetric changes detectable with the methods used in the present study.

Of note, despite observing gross anatomical differences in neuroanatomy and extensive literature supporting a link between pesticides and PD risk (4, 9, 10, 13), we only observed a trending effect of occupational pesticide exposure with increased PD risk. We believe the most likely explanation for the less significant effect compared to previous studies is that our self-reported measure of pesticide exposure is noisy and may not precisely capture exposure to the most neurotoxic OPs. Another possible factor for our conflicting, non-significant finding may be due to our study’s increased sample size relative to previous case-controlled studies investigating the relationship between pesticide exposure and PD diagnosis. One review paper found that only 3 out of 38 case-controlled studies had more than 375 instances of PD diagnosis, with the majority of studies maintaining a sample size of less than or equal to 150 instances (39), whereas our survival analysis consists of 551 instances of PD diagnosis and exposure to pesticides. The UK BioBank occupational pesticide exposure questionnaire captures only the frequency of occupational pesticide exposure but lacks information regarding the dose, duration, or specific chemical composition of pollutants individuals were exposed to. Different pesticides act through distinct neurotoxic mechanisms, and a non-specific exposure measure likely masks these nuances, leading to diffuse volumetric effects across multiple regions (13, 36). Repeated or chronic exposure may also trigger cascading cycles of neuronal damage and atrophy, culminating in widespread structural brain differences. However, this remains speculative and requires further investigation using more precise measures of exposure and neuroimaging techniques capable of capturing microstructural alterations. For the purposes of this study, we utilized non-invasive MRI scans, which are appropriate for detecting global brain effects, but the same cannot be said about cellular mechanisms within regional microstructure.

The most robust associations were negative correlations between pesticide exposure and total gray matter, cortical volume, and thalamic volume, all of which survived multiple comparison corrections. Gray matter volume is a well-established marker of global brain health, with age-related reductions commonly observed in normal aging (40). More pronounced reductions are a characteristic feature of numerous neurodegenerative diseases, including Alzheimer’s disease, PD, Huntington’s disease, and multiple sclerosis (41, 42, 43, 44). Environmental factors such as alcohol consumption and air pollution have also been linked to decreased gray matter volumes (45, 46). Our study, which is substantially larger and population-based, strengthens the evidence that occupational pesticide exposure contributes to volumetric reductions in total gray matter and suggests that these effects may be widespread across both cortical and subcortical regions. We also observed significant bilateral reductions in cortical volume among individuals exposed to pesticides. Cortical gray matter reductions have previously been linked to environmental pollutants in adults (47) and to cortical thickness alterations in children with prenatal exposure to organophosphates, including chlorpyrifos (15, 48, 49, 50). Specifically, high prenatal chlorpyrifos exposure has been associated with enlargement of select cortical regions, such as the superior frontal gyrus, interior postcentral gyrus, and superior temporal gyrus (48). In contrast, our findings in adult participants reveal global cortical volume reductions rather than localized cortical enlargement, suggesting fundamental differences between neurodevelopmental effects of prenatal exposure to pesticides and the neurodegenerative relationship in occupationally exposed adults. Differences in study specifics, such as participant age, exposure timing, and measurement techniques likely contribute to differing results. For example, the mean age of our cohort was 64.49 years, while prior studies examined children aged 6-14.7 years and used region-specific cortical thickness measurements, whereas our study relied mainly on global surface-based voxel counts.

Our third most significant finding was the negative association between pesticide exposure and thalamic volume bilaterally. The thalamus plays a critical role in sensory integration and motor control, and has been implicated as both a structural and functional regional biomarker in PD (51, 53). Neurodevelopmental studies have linked elevated prenatal pesticide metabolite levels to lower thalamic volume in children (15), a finding that parallels our results in adults and suggests that the thalamus may be particularly vulnerable to pesticide-related neurotoxicity across an individual’s lifespan.

Finally, we observed a positive association between occupational pesticide exposure and white matter hyperintensities (WMHs). WMHs are radiological markers of white matter damage seen as hyperintense regions in MRI sequencing often associated with microvascular pathology, neuroinflammation, demyelination and blood-brain barrier disruption (52). WMHs are considered predictive measures of stroke, dementia, and other neurovascular diseases (54). While WMHs are often attributed to small vessel ischemia, emerging evidence suggests that environmental toxicants, including pesticides, may also directly contribute to their development. Case reports have documented toxic leukoencephalopathy following pesticide exposure, with MRI revealing diffuse white matter abnormalities that resolve upon cessation, indicating direct toxic effects on myelinated fibers (54, 55). Population-level studies further link environmental exposures to greater WMH burden while controlling for vascular risk factors (56). Increased prenatal chlorpyrifos exposure was found to be associated with frontal and parietal cortical thinning in children later in life, suggesting a relationship between white matter integrity and environmental pollutants in the developing brain (15). The positive association between pesticide exposure and increased WMH volume further confirms and supports the growing literature surrounding the relationship between neurotoxicants and white matter atrophy. Mechanistically, pesticide exposure may promote WMHs through microvascular injury, blood-brain barrier disruption, oxidative stress, or direct oligodendroglial damage, however longitudinal imaging and biomarker studies are required to further investigate this pathological and clinical significance.

A major limitation of this study is the reliance on a self-report, four-point occupational questionnaire as the primary variable to measure pesticide exposure. This measure lacks detail regarding the type, concentration, and duration of pesticide exposure, all of which are critical determinants of neurotoxicity (57). Despite this limitation, we were able to detect significant, region-specific associations, underscoring the sensitivity of MRI-based volumetric measures. Future research should incorporate objective biomarkers of pesticide exposure, such as blood assays in human adults, and investigate the potential link to structural and functional imaging outcomes. Another limitation is the observational design of the study, which precludes strong causal claims as our observed associations may be the result of an unmeasured confound. To mitigate this we did attempt to control for ancestral and socioeconomic factors, in addition to controlling for differences in cognitive performance in supplementary sensitivity analysis. Despite this, we believe that in the context of previous human and animal studies, our findings support a link in pesticides reducing gray matter volumes and increasing white matter hyperintensities.

Pesticide exposure is only one component of a complex, multifactorial process underlying neurodegeneration. Genetic predispositions, such as mutations in PD-related genes, likely interact with environmental exposures to determine the extent and severity of neuropathological outcomes (57). Other environmental factors, including iron accumulation, air pollution, and dietary patterns, may also influence vulnerability to pesticide-related neurotoxicity (58). Future research should employ integrative approaches that combine genetic, environmental, and neuroimaging techniques to develop comprehensive models of neurodegenerative risk. Such approaches could reveal subgroups of individuals who are particularly susceptible to pesticide exposure and inform targeted prevention strategies. This study provides novel evidence linking occupational pesticide exposure to structural brain differences in adults, including reductions in gray matter, cortical, and thalamic volumes, as well as increases in white matter hyperintensities. These findings suggest that pesticide exposure may contribute to widespread neurodegenerative processes, even in later life. Although the association between pesticide exposure and PD diagnosis did not reach significance, our results align with prior research implicating pesticides as environmental risk factors for PD and other neurodegenerative diseases. Future research should focus on refining exposure assessments to improve accuracy and reliability, incorporate longitudinal imaging, and integrating genetic data to disentangle the complex interplay of factors contributing to neurodegeneration. By advancing our understanding of how environmental toxicants such as pesticides impact the brain, this research has the potential to inform public health interventions aimed at reducing exposure and mitigating adverse neurological disease risk.

## Data Availability

The data used in our analyses was accessed from UK BioBank under accession number 27412. Materials provided by UK BioBank cannot be shared according to UK BioBank's Terms of Use (Section 2.2)

## Supplementary Methods

The average fluid intelligence score (data-field 20016) across all visits was calculated for each individual and included as a confounding variable for further analyses. We wanted to ensure that our significant global brain effects (specifically total gray volume and average cortical volume) were not being driven by the relationship between pesticide exposure confounding with cognitive performance, which is known to also be strongly associated with global brain measures (59, 60). Previous findings have also shown a link between fluid intelligence scores and changes in cortical volume and white matter microstructure, further motivating our inclusion of fluid intelligence scores in the following analyses (61, 62, 63).

In order to investigate lateralization effects among regions located in both hemispheres, we subtracted the right hemisphere region from the left hemisphere region and subsequently labeled the difference of these regions as “ROI_diff” (64). In this operationalization a significant positive effect can be interpreted as a right lateralized effect, for a given region, and a negative effect as a left lateralized effect. All other potential covariates mentioned in the Methods section were included in our supplementary analyses as well.

## Supplementary Results

As shown in Supplementary Figure 1, Supplementary Figure 3 and Supplementary Figure 5, we found there to be no significant association between the difference across hemispheres of specific regions and occupational pesticide exposure. As such, we find there to be no hemispheric lateralization with regards to volumetric differences in brain regions associated with pesticide exposure.

As shown in Supplementary Figure 6, the model estimates of regional volumetric differences when accounting for fluid intelligence scores is similar to the model estimates that do not take into account this variable. Therefore, we can conclude that the link between pesticide exposure and brain morphology is unlikely to be driven by differences in cognitive performance.

## UK BioBank Acknowledgement

This work uses data provided by patients and collected by the NHS as part of their care and support. UK BioBank’s research resource is a major contributor in the advancement of modern medicine and treatment, enabling better understanding of the prevention, diagnosis, and treatment of a wide range of serious and life-threatening illnesses - including cancer, heart disease, and stroke. UK BioBank is generously supported by its founding funders the Wellcome Trust and UK Medical Research Council, as well as the Department of Health, Scottish Government, the Northwest Regional Development Agency, British Heart Foundation, and Cancer Research UK. The organization has over 150 dedicated members of staff, based in multiple locations across the UK. This research used data assets made available by National Safe Haven as part of the Data and Connectivity Core Study, led by Health Data Research UK in partnership with the Office for National Statistics and funded by UK Research and Innovation. This research has been conducted using the UK BioBank Resource under accession number 27412.

## Citations

1) Biundo, R., Weis, L. & Antonini, A. Cognitive decline in Parkinson’s disease: the complex picture. npj Parkinson’s Disease 2, 16018 (2016). 10.1038/npjparkd.2016.18

2) Heijmans, M., Habets, J.G.V., Herff, C. et al. Monitoring Parkinson’s disease symptoms during daily life: a feasibility study. npj Parkinsons Dis. 5, 21 (2019). 10.1038/s41531-019-0093-5

3) Kalia LV, Lang AE. Parkinson’s disease. Lancet. 2015 Aug 29;386(9996):896–912. doi: 10.1016/S0140-6736(14)61393-3. Epub 2015 Apr 19. PMID: 25904081.

4) Warner, T.T. and Schapira, A.H.V. (2003), Genetic and environmental factors in the cause of Parkinson’s disease. Ann Neurol., 53: S16–S25. 10.1002/ana.10487

5) Dickson DW. Neuropathology of Parkinson disease. Parkinsonism Relat Disord. 2018 Jan;46 Suppl 1(Suppl 1):S30–S33. doi: 10.1016/j.parkreldis.2017.07.033. Epub 2017 Aug 1. PMID: 28780180; PMCID: PMC5718208.

6) Braak, H., Braak, E. Pathoanatomy of Parkinson’s disease. J Neurol 247 (Suppl 2), II3–II10 (2000). 10.1007/PL00007758

7) Kaur K, Kaur R. Occupational Pesticide Exposure, Impaired DNA Repair, and Diseases. Indian J Occup Environ Med. 2018 May-Aug;22(2):74–81. doi: 10.4103/ijoem.IJOEM_45_18. PMID: 30319227; PMCID: PMC6176703.

8) Freire C, Koifman S. Pesticide exposure and Parkinson’s disease: epidemiological evidence of association. Neurotoxicology. 2012 Oct;33(5):947–71. doi: 10.1016/j.neuro.2012.05.011. Epub 2012 May 22. PMID: 22627180.

9) Rodrigo Franco, Sumin Li, Humberto Rodriguez-Rocha, Michaela Burns, Mihalis I. Panayiotidis, Molecular mechanisms of pesticide-induced neurotoxicity: Relevance to Parkinson’s disease, Chemico-Biological Interactions, Volume 188, Issue 2, 2010, Pages 289–300, ISSN 0009-2797, 10.1016/j.cbi.2010.06.003.

10) Baltazar MT, Dinis-Oliveira RJ, de Lourdes Bastos M, Tsatsakis AM, Duarte JA, Carvalho F. Pesticides exposure as etiological factors of Parkinson’s disease and other neurodegenerative diseases--a mechanistic approach. Toxicol Lett. 2014 Oct 15;230(2):85–103. doi: 10.1016/j.toxlet.2014.01.039. Epub 2014 Feb 3. PMID: 24503016.

11) Tanner CM, Kamel F, Ross GW, Hoppin JA, Goldman SM, Korell M, Marras C, Bhudhikanok GS, Kasten M, Chade AR, Comyns K, Richards MB, Meng C, Priestley B, Fernandez HH, Cambi F, Umbach DM, Blair A, Sandler DP, Langston JW. Rotenone, paraquat, and Parkinson’s disease. Environ Health Perspect. 2011 Jun;119(6):866–72. doi: 10.1289/ehp.1002839. Epub 2011 Jan 26. PMID: 21269927; PMCID: PMC3114824.

12) Richardson JR, Fitsanakis V, Westerink RHS, Kanthasamy AG. Neurotoxicity of pesticides. Acta Neuropathol. 2019 Sep;138(3):343–362. doi: 10.1007/s00401-019-02033-9. Epub 2019 Jun 13. PMID: 31197504; PMCID: PMC6826260.

13) Paul, K.C., Krolewski, R.C., Lucumi Moreno, E. et al. A pesticide and iPSC dopaminergic neuron screen identifies and classifies Parkinson-relevant pesticides. Nat Commun 14, 2803 (2023). 10.1038/s41467-023-38215-z

14) Rajput AH, Uitti RJ, Stern W, Laverty W, O’Donnell K, O’Donnell D, Yuen WK, Dua A. Geography, drinking water chemistry, pesticides and herbicides and the etiology of Parkinson’s disease. Can J Neurol Sci. 1987 Aug;14(3 Suppl):414–8. doi: 10.1017/s0317167100037823. PMID: 3676917.

15) van den Dries MA, Lamballais S, El Marroun H, Pronk A, Spaan S, Ferguson KK, Longnecker MP, Tiemeier H, Guxens M. Prenatal exposure to organophosphate pesticides and brain morphology and white matter microstructure in preadolescents. Environ Res. 2020 Dec;191:110047. doi: 10.1016/j.envres.2020.110047. Epub 2020 Aug 14. PMID: 32805249; PMCID: PMC7657967.

16) Chao LL, Rothlind JC, Cardenas VA, Meyerhoff DJ, Weiner MW. Effects of low-level exposure to sarin and cyclosarin during the 1991 Gulf War on brain function and brain structure in US veterans. Neurotoxicology. 2010 Sep;31(5):493–501. doi: 10.1016/j.neuro.2010.05.006. Epub 2010 May 24. PMID: 20580739; PMCID: PMC2934883

17) Laurienti PJ, Burdette JH, Talton J, Pope CN, Summers P, Walker FO, Quandt SA, Lyday RG, Chen H, Howard TD, Arcury TA. Brain Anatomy in Latino Farmworkers Exposed to Pesticides and Nicotine. J Occup Environ Med. 2016 May;58(5):436–43. doi: 10.1097/JOM.0000000000000712. PMID: 27158949; PMCID: PMC4866817.

18) Bagdasarov, A., Reuben, A., Gaffrey, M., Fowler, C., & Camacho, N. (2023). Toxicant exposure and the developing brain: A systematic review of the structural and functional MRI literature. Neuroscience and Biobehavioral Reviews, 144, 105006. 10.1016/j.neubiorev.2022.105006

19) González-Alzaga B, Lacasaña M, Aguilar-Garduño C, Rodríguez-Barranco M, Ballester F, Rebagliato M, Hernández AF. A systematic review of neurodevelopmental effects of prenatal and postnatal organophosphate pesticide exposure. Toxicol Lett. 2014 Oct 15;230(2):104–21. doi: 10.1016/j.toxlet.2013.11.019. Epub 2013 Nov 26. PMID: 24291036.

20) Eskenazi B, Rosas LG, Marks AR, Bradman A, Harley K, Holland N, Johnson C, Fenster L, Barr DB. Pesticide toxicity and the developing brain. Basic Clin Pharmacol Toxicol. 2008 Feb;102(2):228–36. doi: 10.1111/j.1742-7843.2007.00171.x. PMID: 18226078.

21) Mullins RJ, Xu S, Pereira EF, Pescrille JD, Todd SW, Mamczarz J, Albuquerque EX, Gullapalli RP. Prenatal exposure of guinea pigs to the organophosphorus pesticide chlorpyrifos disrupts the structural and functional integrity of the brain. Neurotoxicology. 2015 May;48:9–20. doi: 10.1016/j.neuro.2015.02.002. Epub 2015 Feb 19. PMID: 25704171; PMCID: PMC4442734.

22) Loughnan R, Ahern J, Tompkins C, et al. Association of Genetic Variant Linked to Hemochromatosis With Brain Magnetic Resonance Imaging Measures of Iron and Movement Disorders. JAMA Neurol. 2022;79(9):919–928. doi:10.1001/jamaneurol.2022.2030

23) Tom Chambers, Richard Anney, Peter N Taylor, Alexander Teumer, Robin P Peeters, Marco Medici, Xavier Caseras, D Aled Rees, Effects of Thyroid Status on Regional Brain Volumes: A Diagnostic and Genetic Imaging Study in UK Biobank, The Journal of Clinical Endocrinology & Metabolism, Volume 106, Issue 3, March 2021, Pages 688–696, 10.1210/clinem/dgaa903

24)) Alfaro-Almagro F, Jenkinson M, Bangerter NK, et al. Image processing and quality control for the first 10,000 brain imaging datasets from UK Biobank. Neuroimage. 2018;166(166):400–424. doi:10.1016/j.neuroimage.2017.10.034

25) Dale AM, Fischl B, Sereno MI. Cortical surface-based analysis. I. Segmentation and surface reconstruction. Neuroimage. 1999 Feb;9(2):179–94. doi: 10.1006/nimg.1998.0395. PMID: 9931268.

26) Fischl B, Salat DH, Busa E, Albert M, Dieterich M, Haselgrove C, van der Kouwe A, Killiany R, Kennedy D, Klaveness S, Montillo A, Makris N, Rosen B, Dale AM. Whole brain segmentation: automated labeling of neuroanatomical structures in the human brain. Neuron. 2002 Jan 31;33(3):341–55. doi: 10.1016/s0896-6273(02)00569-x. PMID: 11832223.

27) Postelnicu G, Zollei L, Fischl B. Combined volumetric and surface registration. IEEE Trans Med Imaging. 2009 Apr;28(4):508–22. doi: 10.1109/TMI.2008.2004426. Epub 2008 Aug 15. PMID: 19273000; PMCID: PMC2761957.

28) Fischl B. FreeSurfer. Neuroimage. 2012 Aug 15;62(2):774–81. doi: 10.1016/j.neuroimage.2012.01.021. Epub 2012 Jan 10. PMID: 22248573; PMCID: PMC3685476.

29) Benjamini, Yoav & Hochberg, Yosef. (1995). Controlling The False Discovery Rate - A Practical And Powerful Approach To Multiple Testing. J. Royal Statist. Soc., Series B. 57. 289–300. 10.2307/2346101.

30) Dunn, O. J. (1961). Multiple Comparisons among Means. Journal of the American Statistical Association, 56(293), 52–64. 10.1080/01621459.1961.10482090

31) Sánchez-Santed F, Colomina MT, Herrero Hernández E. Organophosphate pesticide exposure and neurodegeneration. Cortex. 2016 Jan;74:417–26. doi: 10.1016/j.cortex.2015.10.003. Epub 2015 Oct 20. PMID: 26687930.

32) Vellingiri B, Chandrasekhar M, Sri Sabari S, Gopalakrishnan AV, Narayanasamy A, Venkatesan D, Iyer M, Kesari K, Dey A. Neurotoxicity of pesticides - A link to neurodegeneration. Ecotoxicol Environ Saf. 2022 Sep 15;243:113972. doi: 10.1016/j.ecoenv.2022.113972. Epub 2022 Aug 24. PMID: 36029574.

33) Peng J, Stevenson FF, Oo ML, Andersen JK. Iron-enhanced paraquat-mediated dopaminergic cell death due to increased oxidative stress as a consequence of microglial activation. Free Radic Biol Med. 2009 Jan 15;46(2):312–20. doi: 10.1016/j.freeradbiomed.2008.10.045. Epub 2008 Nov 7. PMID: 19027846; PMCID: PMC2654268.

34) Lushchak VI, Matviishyn TM, Husak VV, Storey JM, Storey KB. Pesticide toxicity: a mechanistic approach. EXCLI J. 2018 Nov 8;17:1101–1136. doi: 10.17179/excli2018-1710. PMID: 30564086; PMCID: PMC6295629.

35) Sule RO, Condon L, Gomes AV. A Common Feature of Pesticides: Oxidative Stress-The Role of Oxidative Stress in Pesticide-Induced Toxicity. Oxid Med Cell Longev. 2022 Jan 19;2022:5563759. doi: 10.1155/2022/5563759. PMID: 35096268; PMCID: PMC8791758.

36) Gupta, R. C. (2004). Brain Regional Heterogeneity and Toxicological Mechanisms of Organophosphates and Carbamates. Toxicology Mechanisms and Methods, 14(3), 103–143. 10.1080/15376520490429175

37) Heusinkveld HJ, Westerink RHS. Comparison of different in vitro cell models for the assessment of pesticide-induced dopaminergic neurotoxicity. Toxicol In Vitro. 2017 Dec;45(Pt 1):81–88. doi: 10.1016/j.tiv.2017.07.030. Epub 2017 Jul 31. PMID: 28774849.

38) McCormack AL, Atienza JG, Johnston LC, Andersen JK, Vu S, Di Monte DA. Role of oxidative stress in paraquat-induced dopaminergic cell degeneration. J Neurochem. 2005 May;93(4):1030–7. doi: 10.1111/j.1471-4159.2005.03088.x. PMID: 15857406.

39) Brown TP, Rumsby PC, Capleton AC, Rushton L, Levy LS. Pesticides and Parkinson’s disease--is there a link? Environ Health Perspect. 2006 Feb;114(2):156–64. doi: 10.1289/ehp.8095. PMID: 16451848; PMCID: PMC1367825.

40) Ge Y, Grossman RI, Babb JS, Rabin ML, Mannon LJ, Kolson DL. Age-related total gray matter and white matter changes in normal adult brain. Part I: volumetric MR imaging analysis. AJNR Am J Neuroradiol. 2002 Sep;23(8):1327–33. PMID: 12223373; PMCID: PMC7976241.

41) Thompson PM, Hayashi KM, de Zubicaray G, Janke AL, Rose SE, Semple J, Herman D, Hong MS, Dittmer SS, Doddrell DM, Toga AW. Dynamics of gray matter loss in Alzheimer’s disease. J Neurosci. 2003 Feb 1;23(3):994–1005. doi: 10.1523/JNEUROSCI.23-03-00994.2003. PMID: 12574429; PMCID: PMC6741905.

42) Melzer TR, Watts R, MacAskill MR, Pitcher TL, Livingston L, Keenan RJ, Dalrymple-Alford JC, Anderson TJ. Grey matter atrophy in cognitively impaired Parkinson’s disease. J Neurol Neurosurg Psychiatry. 2012 Feb;83(2):188–94. doi: 10.1136/jnnp-2011-300828. Epub 2011 Sep 2. PMID: 21890574.

43) Halliday GM, McRitchie DA, Macdonald V, Double KL, Trent RJ, McCusker E. Regional specificity of brain atrophy in Huntington’s disease. Exp Neurol. 1998 Dec;154(2):663–72. doi: 10.1006/exnr.1998.6919. PMID: 9878201.

44) Sastre-Garriga J, Ingle GT, Chard DT, Cercignani M, Ramió-Torrentà L, Miller DH, Thompson AJ. Grey and white matter volume changes in early primary progressive multiple sclerosis: a longitudinal study. Brain. 2005 Jun;128(Pt 6):1454–60. doi: 10.1093/brain/awh498. Epub 2005 Apr 7. PMID: 15817511.

45) Daviet R, Aydogan G, Jagannathan K, Spilka N, Koellinger PD, Kranzler HR, Nave G, Wetherill RR. Associations between alcohol consumption and gray and white matter volumes in the UK Biobank. Nat Commun. 2022 Mar 4;13(1):1175. doi: 10.1038/s41467-022-28735-5. PMID: 35246521; PMCID: PMC8897479.

46) Erickson LD, Gale SD, Anderson JE, Brown BL, Hedges DW. Association between Exposure to Air Pollution and Total Gray Matter and Total White Matter Volumes in Adults: A Cross-Sectional Study. Brain Sci. 2020 Mar 13;10(3):164. doi: 10.3390/brainsci10030164. PMID: 32182984; PMCID: PMC7139378.

47) Vu HT, Pham TN, Yokawa T, Nishijo M, The TP, Do Q, Nishino Y, Nishijo H. Alterations in Regional Brain Regional Volume Associated with Dioxin Exposure in Men Living in the Most Dioxin-Contaminated Area in Vietnam: Magnetic Resonance Imaging (MRI) Analysis Using Voxel-Based Morphometry (VBM). Toxics. 2021 Dec 15;9(12):353. doi: 10.3390/toxics9120353. PMID: 34941787; PMCID: PMC8703540.

48) Rauh VA, Perera FP, Horton MK, Whyatt RM, Bansal R, Hao X, Liu J, Barr DB, Slotkin TA, Peterson BS. Brain anomalies in children exposed prenatally to a common organophosphate pesticide. Proc Natl Acad Sci U S A. 2012 May 15;109(20):7871–6. doi: 10.1073/pnas.1203396109. Epub 2012 Apr 30. PMID: 22547821; PMCID: PMC3356641.

49) Sagiv SK, Baker JM, Rauch S, Gao Y, Gunier RB, Mora AM, Kogut K, Bradman A, Eskenazi B, Reiss AL. Prenatal and childhood exposure to organophosphate pesticides and functional brain imaging in young adults. Environ Res. 2024 Feb 1;242:117756. doi: 10.1016/j.envres.2023.117756. Epub 2023 Nov 26. PMID: 38016496; PMCID: PMC11298288.

50) Peterson BS, Delavari S, Bansal R, Sawardekar S, Gupte C, Andrews H, Hoepner LA, Garcia W, Perera F, Rauh V. Brain Abnormalities in Children Exposed Prenatally to the Pesticide Chlorpyrifos. JAMA Neurol. 2025 Aug 18;82(10):1057–68. doi: 10.1001/jamaneurol.2025.2818. Epub ahead of print. PMID: 40824645; PMCID: PMC12362277.

51) Younce JR, Campbell MC, Perlmutter JS, Norris SA. Thalamic and ventricular volumes predict motor response to deep brain stimulation for Parkinson’s disease. Parkinsonism Relat Disord. 2019 Apr;61:64–69. doi: 10.1016/j.parkreldis.2018.11.026. Epub 2018 Nov 28. PMID: 30527905; PMCID: PMC6488428.

52) Ottavi TP, Pepper E, Bateman G, Fiorentino M, Brodtmann A. Consensus statement for the management of incidentally found brain white matter hyperintensities in general medical practice. Med J Aust. 2023 Sep 18;219(6):278–284. doi: 10.5694/mja2.52079. Epub 2023 Aug 21. PMID: 37604652.

53) Lee SH, Kim SS, Tae WS, Lee SY, Choi JW, Koh SB, Kwon DY. Regional volume analysis of the Parkinson disease brain in early disease stage: gray matter, white matter, striatum, and thalamus. AJNR Am J Neuroradiol. 2011 Apr;32(4):682–7. doi: 10.3174/ajnr.A2372. Epub 2011 Feb 17. PMID: 21330396; PMCID: PMC7965874.

54) Debette S, Markus HS. The clinical importance of white matter hyperintensities on brain magnetic resonance imaging: systematic review and meta-analysis. BMJ. 2010 Jul 26;341:c3666. doi: 10.1136/bmj.c3666. PMID: 20660506; PMCID: PMC2910261.

55) Baek BH, Kim SK, Yoon W, Heo TW, Lee YY, Kang HK. Chlorfenapyr-Induced Toxic Leukoencephalopathy with Radiologic Reversibility: A Case Report and Literature Review. Korean J Radiol. 2016 Mar-Apr;17(2):277–80. doi: 10.3348/kjr.2016.17.2.277. Epub 2016 Mar 2. PMID: 26957914; PMCID: PMC4781768.

56) Schoemaker D, Zanon Zotin MC, Chen K, Igwe KC, Vila-Castelar C, Martinez J, Baena A, Fox-Fuller JT, Lopera F, Reiman EM, Brickman AM, Quiroz YT. White matter hyperintensities are a prominent feature of autosomal dominant Alzheimer’s disease that emerge prior to dementia. Alzheimers Res Ther. 2022 Jun 29;14(1):89. doi: 10.1186/s13195-022-01030-7. PMID: 35768838; PMCID: PMC9245224.

57) Peng J, Peng L, Stevenson FF, Doctrow SR, Andersen JK. Iron and paraquat as synergistic environmental risk factors in sporadic Parkinson’s disease accelerate age-related neurodegeneration. J Neurosci. 2007 Jun 27;27(26):6914–22. doi: 10.1523/JNEUROSCI.1569-07.2007. PMID: 17596439; PMCID: PMC6672233.

58)) Dietary Factors Affect Brain Iron Accumulation and Parkinson’s Disease Risk Jonathan Ahern, Mary ET Boyle, Leo Sugrue, Ole Andreassen, Anders Dale, Wesley K. Thompson, Chun Chieh Fan, Robert Loughnan medRxiv 2024.03.13.24304253; doi: 10.1101/2024.03.13.24304253

59) Chen, Pin-Yu, et al. “Fluid intelligence is associated with cortical volume and white matter tract integrity within multiple-demand system across adult lifespan.” NeuroImage 212 (2020): 116576. ISSN 1053-8119, DOI: 10.1016/j.neuroimage.2020.116576.

60) DeCarli C, Murphy DG, Tranh M, Grady CL, Haxby JV, Gillette JA, Salerno JA, Gonzales-Aviles A, Horwitz B, Rapoport SI, et al. The effect of white matter hyperintensity volume on brain structure, cognitive performance, and cerebral metabolism of glucose in 51 healthy adults. Neurology. 1995 Nov;45(11):2077–84. doi: 10.1212/wnl.45.11.2077. PMID: 7501162.

61)) Coupled Changes in Brain White Matter Microstructure and Fluid Intelligence in Later Life Stuart J. Ritchie, Mark E. Bastin, Elliot M. Tucker-Drob, Susana Muñoz Maniega, Laura E. Engelhardt, Simon R. Cox, Natalie A. Royle, Alan J. Gow, Janie Corley, Alison Pattie, Adele M. Taylor, Maria del C. Valdés Hernández, John M Starr, Joanna M. Wardlaw, Ian J. Deary, Journal of Neuroscience 3 June 2015, 35 (22) 8672–8682; DOI: 10.1523/JNEUROSCI.0862-15.2015

62) Rabbitt P, Scott M, Thacker N, Lowe C, Jackson A, Horan M, Pendleton N. Losses in gross brain volume and cerebral blood flow account for age-related differences in speed but not in fluid intelligence. Neuropsychology. 2006 Sep;20(5):549–557. doi: 10.1037/0894-4105.20.5.549. PMID: 16938017.

63) Raz N, Lindenberger U, Ghisletta P, Rodrigue KM, Kennedy KM, Acker JD. Neuroanatomical correlates of fluid intelligence in healthy adults and persons with vascular risk factors. Cereb Cortex. 2008 Mar;18(3):718–26. doi: 10.1093/cercor/bhm108. Epub 2007 Jul 5. PMID: 17615248; PMCID: PMC2657233.

64) Riederer P, Sian-Hülsmann J. The significance of neuronal lateralisation in Parkinson’s disease. J Neural Transm (Vienna). 2012 Aug;119(8):953–62. doi: 10.1007/s00702-012-0775-1. Epub 2012 Feb 26. PMID: 22367437.

