## Supplement 1 for "Occupational Pesticide Exposure and Regional Brain Volume Differences in UK BioBank"

### Supplementary Figures & Tables

#### Supplementary Table 1: ROI Field Descriptions

| Coded ROI Label (as seen in Figures) | FreeSurfer ROI Label | Region Description |
|---|---|---|
| LatVentVol_lh | Lateral Ventricle - Left Hemisphere | CSF-filled cavity in the left cerebral hemisphere, including frontal, temporal, occipital horns and body. |
| VentricleChoroidVol | Ventricular and Choroid Plexus Volume | Combined volume of all ventricular CSF including lateral, inferior lateral, third, fourth, and fifth ventricles, together with the choroid plexus |
| CerebellumCortexVol_lh | Cerebellum Cortex - Left Hemisphere | Gray matter mantle of the left cerebellum (folia), excluding cerebellum white matter and brainstem |
| CerebellumWhiteMatterVol_lh | Cerebellum White Matter - Left Hemisphere | Myelinated cerebellar tissue (arbor vitae) in the left cerebellar hemisphere, excluding cerebellar cortex and brainstem |
| EstimatedTotalIntraCranialVol | Total Intracranial Volume (ICV) | Estimate of the intracranial volume based on the talairach transform. This is the same measure as Estimated Total Intracranial Volume (eTIV). |
| BrainStem | Brainstem | Midbrain, pons, and medulla oblongata, excluding ventral diencephalon and cerebellum |
| totalGrayVol | Total Gray Matter | Total gray matter volume. This is simply the sum of lhCortex + rhCortex + SubCortGray + CerebellumGM. As such, it includes both surface-based volume calculations and voxel counts. |
| VesselVol_rh | Intracranial Vessels - Right Hemisphere | Voxels labeled as intracranial vessels in the right hemisphere (e.g., large cortical vessels/dural sinuses adjacent to cortex). Not neural tissue. |
| PutamenVol_lh | Putamen - Left Hemisphere | Left lenticular basal ganglia nucleus involved in motor planning and learning; lateral to globus pallidus and medial to external capsule |
| SubCortGrayVol | Sub Cortical Gray Matter | Includes thalamus, caudate, putamen, pallidum, hippocampus, amygdala, accumbens, ventral DC, substantia nigra (if there). This is a simple voxel count of structures identified as subcortical GM. |
| HippocampVol_rh | Hippocampus - Right Hemisphere | Right medial temporal lobe structures including the head, body, and tail, which are essential for episodic memory; excludes adjacent choroid plexus |
| PutamenVol_rh | Putamen - Right Hemisphere | Right lenticular basal ganglia nucleus essential for motor control and habit learning; lateral to globus pallidus |

| | | |
|---|---|---|
| HippocampVol_lh | Hippocampus - Left Hemisphere | Left medial temporal lobe hippocampal formation including the head, body, and tail; essential for memory encoding/navigation; excludes choroid plexus |
| AmygdalaVol_rh | Amygdala - Right Hemisphere | Right basolateral/centromedial amygdaloid complex in the anterior medial temporal lobe; affective/associative processing |
| BrainSegNotVentSurfVol | BrainSegNotVent | Volume estimated using surface-based ribbon volumes where applicable; includes cerebrum and cerebellum but excludes ventricles, CSF, choroid plexus, and vessels. |
| BrainSegNotVentVol | BrainSegNotVent | Sum of the volume of the structures identified in the aseg.mgz volume. This will include cerebellum but not ventricles, CSF, and dura. Includes partial volume compensation. |
| CerebellumWhiteMatterVol_rh | Cerebellum White Matter - Right Hemisphere | Cerebral (supratentorial) right hemisphere white matter within the white surface; includes WM hypointensities, excludes cerebellum and brainstem |
| ChoroidPlexusVol_rh | Choroid Plexus Volume - Right Hemisphere | Vascular epithelium within the ventricular system, primarily the lateral ventricle body and temporal horn, producing CSF; right side |
| CC_Mid_Anterior | Mid Anterior Cingulate Cortex | Second fifth (from the genu posteriorly) of the midsagittal corpus callosum partition; connects the rostral anterior frontal and anterior cingulate cortex regions |
| Cortex_rh | Cortex - Right Hemisphere | Volume inside the pial surface minus the volume inside the white surface minus tissue inside the ribbon that is not part of cortex (eg, hippocampus). This uses the surface-based volume calculation and is not a voxel count. It is approximately equal to the count of cortical label voxels |
| CSF | Cerebral Spinal Fluid (CSF) | Intracranial CSF volume including the ventricular and subarachnoid spaces, identified on T1-weighted segmentation |
| SupraTentorialVolNotVent | Non-Ventricle Supratentorial | Same as SupraTentorial but subtracting the volume ventricles (lateral,inferior lateral, 3rd, 4th, 5th), CSF, and choroid plexus. |
| ChoroidPlexusVol_lh | Choroid Plexus Volume - Left Hemisphere | Vascular epithelium within the ventricular system, primarily the lateral ventricle body and temporal horn, producing CSF; left side |
| FourthVent | 4th Ventricle | Volumetric segmentation of the fourth ventricle |
| CaudateVol_rh | Caudate - Right Hemisphere | Right dorsal striatal nucleus including the head, body, and tail, bordering the lateral ventricle; involved in cognitive and motor circuitry |
| InfLatVentVol_rh | Inferior Lateral Ventricle - Right Hemisphere | CSF in the right temporal/inferior horn of the lateral ventricle |
| OpticChiasm | Optic Chiasm | Decussation of retinal ganglion cell axons at the base of the skull; part of the anterior visual pathway |
| AccumbensVol_lh | Accumbens - Left Hemisphere | Left ventral striatal nucleus at the caudate-putamen-septal junction; involved in reward and motivation processing |
| FifthVent | 5th Ventricle | Volumetric segmentation of the fifth ventricle. Referred to as "cavum septum pallucidum", and is regarded by FreeSurfer as a "bit of an enigma that tends to only occur on occasion". Although it is not reported in any |

| | | |
|---|---|---|
| | | manuscripts, it was confirmed by neuroradiologist, P. Ellen Grant |
| CerebralWhiteMatterVol_rh | Cerebral White Matter - Right Hemisphere | Volume inside the white surface minus anything that is not WM. CerebralWhiteMatter includes hypointensities. Does not include cerebellar white matter or brainstem. This uses the surface-based volume computation for part of the calculation and counts voxels to subtract "anything not in WM". It is approximately equal to the count of WM voxels |
| SupraTentorialVol | Supratentorial | Includes everything except cerebellum (GM and WM) and brain stem. It is computed based on everything inside the pial surface plus any structures that might fall partially or totally outside of the pial, e.g., hippocampus, amygdala, corpus callosum, ventral DC, thalamus, ventricles, and choroid plexus. The function that determines the volume of structures outside of the supratentorium is SupraTentorialVolCorrection() in cma.c. Note that SupraTentorial includes ventricle, choroid plexus, and vessel. This uses surface-based volume calculations and voxel counts |
| VesselVol_lh | Intracranial Vessels - Left Hemisphere | Voxels labeled as intracranial vessels in the left hemisphere (large cortical/dural vessels). Not neural tissue. |
| InfLatVentVol_lh | Inferior Lateral Ventricle - Left Hemisphere | CSF in the left temporal (inferior) horn of the lateral ventricle |
| AmygdalaVol_lh | Amygdala - Left Hemisphere | Left amygdaloid complex in the anterior medial temporal lobe; related to emotion, salience, and associative learning |
| ThalamusVol_rh | Thalamus - Right Hemisphere | With regards to the right hemisphere, the thalamus proper (including all thalamic nuclei except the lateral and medial geniculate bodies) was defined on all coronal slices on which it was present, from the first slice containing the anterior nucleus through the last slice containing the pulvinar nucleus. The structure was bounded medially by the third ventricle, and laterally by the internal capsule. The superior border was the body of the lateral ventricle, and the inferior border, the hypothalamic fissure. |
| WMhyperintensities | White Matter Hyperintensities | Global white matter tracts, lesions/hypointense clusters on T1 within the white-matter mask (often small-vessel disease/WM lesions); labeled as “WM-hypointensities” |
| BrainSegVol_to_eTIV | | Ratio of BrainSegVol to Estimated Total Intracranial Volume (eTIV); a head-size normalized brain volume measure |
| CC_Central | Central Cingulate Cortex | Middle fifth of the midsagittal corpus callosum partition; interconnects premotor/motor/somatosensory cortices |
| LatVentVol_rh | Lateral Ventricle - Right Hemisphere | CSF-filled cavity in the right cerebral hemisphere (frontal, temporal, and occipital horns including the body). |
| CaudateVol_lh | Caudate - Left Hemisphere | Left dorsal striatal nucleus, including head, body, and tail, bordering the lateral ventricle; involved in cognitive and motor circuitry |
| VentralDCVol_rh | Ventral Diencephalon - Right Hemisphere | With respect to the right hemisphere, this region consists of volumetric segmentation containing the hypothalamus, basal forebrain, and sublenticular extended amygdala (SLEA), as well as a large portion of ventral tegmentum (which is also included in our ventral diencephalon |

| | | |
|---|---|---|
| | | region by convention although part of the midbrain) |
| CC_Posterior | Posterior Cingulate Cortex | Posterior fifth (splenial region) of midsagittal corpus callosum; fibers to parietal, occipital, and temporal association cortices |
| MaskVol_to_eTIV | | Ratio of brainmask.mgz volume (union of surfaces and aseg labels approximating intracranial brain compartment) to eTIV |
| PallidumVol_rh | Pallidum - Right Hemisphere | Right globus pallidus, including internal and external segments, medial to putamen; motor |
| Cortex_lh | Cortex - Left Hemisphere | Volume inside the pial surface minus the volume inside the white surface minus tissue inside the ribbon that is not part of cortex (eg, hippocampus). This uses the surface-based volume calculation and is not a voxel count. It is approximately equal to the count of cortical label voxels |
| CerebralWhiteMatterVol_lh | Cerebral White Matter - Left Hemisphere | Volume inside the white surface minus anything that is not WM. CerebralWhiteMatter includes hypointensities. Does not include cerebellar white matter or brainstem. This uses the surface-based volume computation for part of the calculation and counts voxels to subtract "anything not in WM". It is approximately equal to the count of WM voxels |
| ThalamusVol_lh | Thalamus - Left Hemisphere | With regards to the left hemisphere, the thalamus proper (including all thalamic nuclei except the lateral and medial geniculate bodies) was defined on all coronal slices on which it was present, from the first slice containing the anterior nucleus through the last slice containing the pulvinar nucleus. The structure was bounded medially by the third ventricle, and laterally by the internal capsule. The superior border was the body of the lateral ventricle, and the inferior border, the hypothalamic fissure. |
| VentralDCVol_lh | Ventral Diencephalon - Left Hemisphere | With respect to the left hemisphere, this region consists of volumetric segmentation containing the hypothalamus, basal forebrain, and sublenticular extended amygdala (SLEA), as well as a large portion of ventral tegmentum (which is also included in our ventral diencephalon region by convention although part of the midbrain) |
| CC_Mid_Posterior | Mid Posterior Cingulate Cortex | Fourth fifth of the midsagittal corpus callosum partition anterior to the splenium. Fibers to the posterior cingulate/parietal areas. |
| AccumbensVol_rh | Nucleus Accumbens - Right Hemisphere | Right ventral striatal nucleus at the caudate-putamen-septal interface; involved in reward and valuation circuitry. |
| CerebellumCortexVol_rh | Cerebellum Cortex - Right Hemisphere | Gray-matter mantle of the right cerebellum (folia), excluding cerebellar white matter and brainstem. |
| non_WMhyperintensities | Non-White Matter Hyperintensities | Hypointense clusters on T1 outside the white-matter mask, often in perivascular spaces and cavities |
| PallidumVol_lh | Pallidum - Left Hemisphere | Left global pallidus, including internal and external, medial to putamen; involves motor output nucleus of basal ganglia |
| ThirdVent | 3rd Ventricle | Volumetric segmentation of the third ventricle |
| CC_Anterior | Anterior Cingulate Cortex | Most anterior fifth (genu region) of the midsagittal corpus callosum; interconnects prefrontal/anterior cingulate regions |
| BrainSegVol | BrainSeg/BrainSegVol | Sum of the volume of the following structures: cerebellum and ventricles |

| | | |
|---|---|---|
| | | but should exclude dura. This does not include partial volume compensation. |

***Supplementary Table 1:*** *Table highlighting coded region of interest (ROI) labels as denoted in Figures, the corresponding labels from FreeSurfer, and descriptions of the region within the human brain itself.*

**<u>Supp. Figure 1:</u> The association between occupational pesticide exposure and average regional brain volume differences**

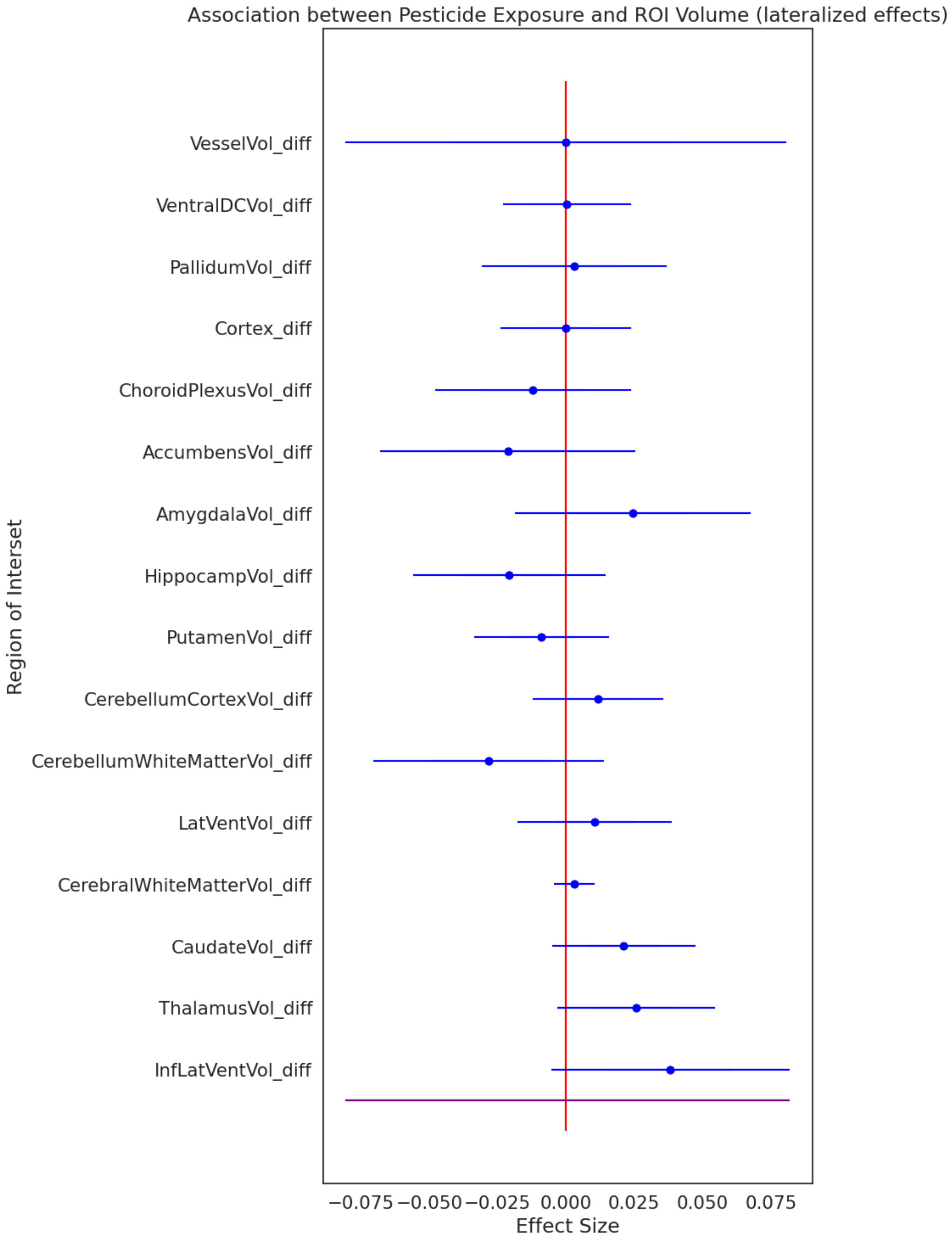

***Supplementary Figure 1:*** *Forest plot depicting the association between occupational pesticide exposure and ROI volume differences using generalized linear regression models. ROI volume differences were calculated by subtracting the right hemisphere ROI from the left hemisphere ROI. As such, a positive effect size corresponds to a right hemisphere lateralized effect and a negative effect size corresponds to a left hemisphere lateralized effect. Standard covariates were used including sex assigned at birth, age at the time of imaging, top 10 principal components of genetic ancestry (C1-C10), Indices of Multiple Deprivation (health, education, and income), clinic site where imaging was conducted, and estimated total intracranial volume.*

#### Supplementary Table 2: Corresponding Model Summary for Supp. Figure 1

| Brain Region | Coef | Standard Errors | CI (low) | CI (high) | N | p_value | p_FDR | p_Bonferroni |
|---|---|---|---|---|---|---|---|---|
| InfLatVentVol_diff | 0.038265 | 0.022315 | -0.005472 | 0.082002 | 21049 | 8.64E-02 | 6.13E-01 | 1 |
| ThalamusVol_diff | 0.025591 | 0.014727 | -0.003273 | 0.054454 | 21049 | 8.23E-02 | 6.13E-01 | 1 |
| CaudateVol_diff | 0.021163 | 0.013424 | -0.005148 | 0.047474 | 21049 | 1.15E-01 | 6.13E-01 | 1 |
| CerebralWhiteMatterVol_diff | 0.003075 | 0.003852 | -0.004474 | 0.010624 | 21049 | 4.25E-01 | 6.80E-01 | 1 |
| LatVentVol_diff | 0.010582 | 0.014373 | -0.017589 | 0.038753 | 21049 | 4.62E-01 | 6.80E-01 | 1 |
| CerebellumWhiteMatterVol_diff | -0.028248 | 0.021484 | -0.070356 | 0.013861 | 21049 | 1.89E-01 | 6.80E-01 | 1 |
| CerebellumCortexVol_diff | 0.011743 | 0.012256 | -0.012279 | 0.035765 | 21049 | 3.38E-01 | 6.80E-01 | 1 |
| PutamenVol_diff | -0.008924 | 0.01257 | -0.033561 | 0.015712 | 21049 | 4.78E-01 | 6.80E-01 | 1 |
| HippocampVol_diff | -0.020739 | 0.018017 | -0.056052 | 0.014574 | 21049 | 2.50E-01 | 6.80E-01 | 1 |
| AmygdalaVol_diff | 0.024501 | 0.021973 | -0.018565 | 0.067567 | 21049 | 2.65E-01 | 6.80E-01 | 1 |
| AccumbensVol_diff | -0.021238 | 0.023867 | -0.068016 | 0.025539 | 21049 | 3.74E-01 | 6.80E-01 | 1 |
| ChoroidPlexusVol_diff | -0.012037 | 0.018285 | -0.047875 | 0.023801 | 21049 | 5.10E-01 | 6.80E-01 | 1 |
| Cortex_diff | -0.000037 | 0.012223 | -0.023993 | 0.023919 | 21049 | 9.98E-01 | 9.99E-01 | 1 |
| PallidumVol_diff | 0.003087 | 0.017298 | -0.030815 | 0.03699 | 21049 | 8.58E-01 | 9.99E-01 | 1 |
| VentralDCVol_diff | 0.000342 | 0.011929 | -0.023039 | 0.023722 | 21049 | 9.77E-01 | 9.99E-01 | 1 |
| VesselVol_diff | -0.000036 | 0.041105 | -0.080599 | 0.080528 | 21049 | 9.99E-01 | 9.99E-01 | 1 |

***Supplementary Table 2:*** *Model summaries corresponding to the Supplementary Figure 1 forest plot. Model summaries include the brain regions of interest (coded ROIs), model estimates*

*(Coef), standard errors, low and high confidence intervals, the sample size (N), the nominal p-value, and the p-values of both FDR and Bonferroni multiple comparisons test for each ROI.*

**<u>Supp. Figure 2:</u> The association between occupational pesticide exposure and average regional brain volume differences (including fluid intelligence scores as a covariate)**

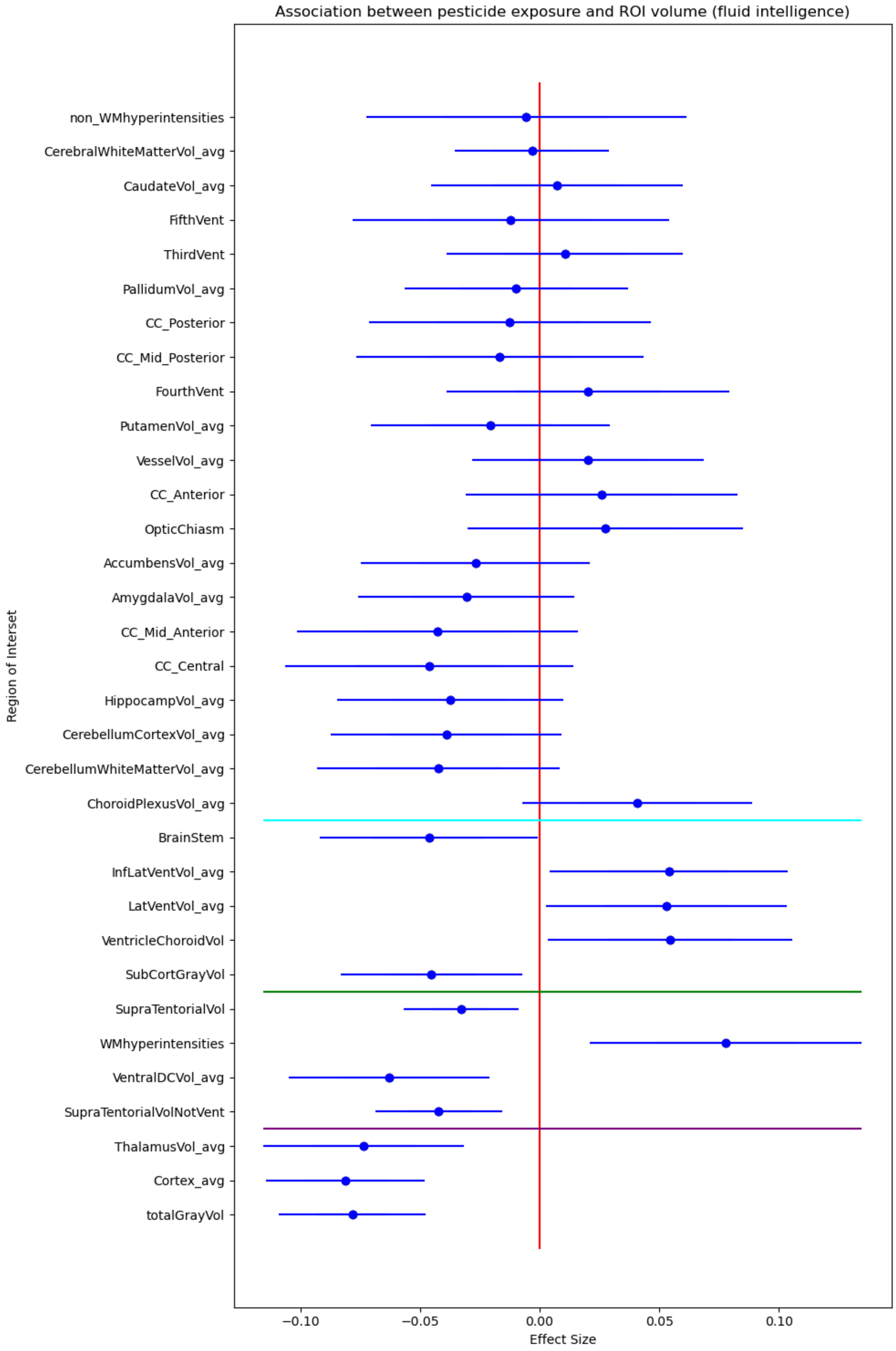

***Supplementary Figure 2:*** *Forest plot depicting the association between occupational pesticide exposure and average regional brain volumes (ROI) using generalized linear regression models. ROI volume values were averaged between hemispheres to account for bilateral effects. Standard covariates were taken into account, including sex assigned at birth, age at the time of imaging, top 10 principal components of genetic ancestry (C1-C10), average fluid intelligence scores (across all visits), clinic site where imaging was conducted, and estimated total intracranial volume. The horizontal blue line represents nominal statistical significance, the horizontal green line represents the significance threshold for False Discovery Rate (FDR) correction, and the horizontal purple line represents the significance threshold for Bonferroni correction.*

#### Supplementary Table 3: Corresponding Model Summary for Supp. Figure 2

| Brain Region | Coef | Standard Errors | CI (low) | CI (high) | N | p_value | p_FDR | p_Bonferroni |
|---|---|---|---|---|---|---|---|---|
| totalGrayVol | -0.078268 | 0.015632 | -0.108906 | -0.04763 | 22542 | 5.53E-07 | 1.80E-05 | 1.80E-05 |
| Cortex_avg | -0.081141 | 0.016891 | -0.114246 | -0.048036 | 22542 | 1.56E-06 | 2.60E-05 | 5.10E-05 |
| ThalamusVol_avg | -0.073491 | 0.021354 | -0.115343 | -0.031639 | 22542 | 5.78E-04 | 6.36E-03 | 1.91E-02 |
| SupraTentorialVolNotVent | -0.042217 | 0.013564 | -0.068803 | -0.015632 | 22542 | 1.86E-03 | 1.53E-02 | 6.12E-02 |
| VentralDCVol_avg | -0.063057 | 0.021397 | -0.104994 | -0.021119 | 22542 | 3.21E-03 | 2.12E-02 | 1.06E-01 |
| WMhyperintensities | 0.077884 | 0.028973 | 0.021098 | 0.13467 | 22542 | 7.18E-03 | 3.49E-02 | 2.37E-01 |
| SupraTentorialVol | -0.03288 | 0.012278 | -0.056945 | -0.008814 | 22542 | 7.41E-03 | 3.49E-02 | 2.45E-01 |
| SubCortGrayVol | -0.045199 | 0.019413 | -0.083247 | -0.007151 | 22542 | 1.99E-02 | 8.21E-02 | 6.57E-01 |
| VentricleChoroidVol | 0.054471 | 0.026033 | 0.003447 | 0.105495 | 22542 | 3.64E-02 | 1.16E-01 | 1.00E+00 |
| LatVentVol_avg | 0.052992 | 0.025634 | 0.002752 | 0.103233 | 22542 | 3.87E-02 | 1.16E-01 | 1.00E+00 |
| InfLatVentVol_avg | 0.054112 | 0.025381 | 0.004366 | 0.103857 | 22542 | 3.30E-02 | 1.16E-01 | 1.00E+00 |
| BrainStem | -0.046273 | 0.023301 | -0.091942 | -0.000604 | 22542 | 4.70E-02 | 1.29E-01 | 1.00E+00 |
| ChoroidPlexusVol_avg | 0.040823 | 0.024529 | -0.007252 | 0.088898 | 22542 | 9.61E-02 | 2.43E-01 | 1.00E+00 |
| CerebellumWhiteMatterVol_avg | -0.042251 | 0.025918 | -0.093049 | 0.008548 | 22542 | 1.03E-01 | 2.43E-01 | 1.00E+00 |
| CerebellumCortexVol_avg | -0.039038 | 0.024668 | -0.087387 | 0.00931 | 22542 | 1.14E-01 | 2.50E-01 | 1.00E+00 |
| HippocampVol_avg | -0.037337 | 0.024155 | -0.084681 | 0.010006 | 22542 | 1.22E-01 | 2.52E-01 | 1.00E+00 |
| CC_Central | -0.046019 | 0.030695 | -0.10618 | 0.014143 | 22542 | 1.34E-01 | 2.60E-01 | 1.00E+00 |
| CC_Mid_Anterior | -0.042668 | 0.029961 | -0.10139 | 0.016054 | 22542 | 1.54E-01 | 2.83E-01 | 1.00E+00 |
| AmygdalaVol_avg | -0.030636 | 0.022992 | -0.0757 | 0.014428 | 22542 | 1.83E-01 | 3.17E-01 | 1.00E+00 |
| AccumbensVol_avg | -0.026847 | 0.024407 | -0.074683 | 0.020989 | 22542 | 2.71E-01 | 4.48E-01 | 1.00E+00 |

| | | | | | | | | |
|---|---|---|---|---|---|---|---|---|
| OpticChiasm | 0.027629 | 0.029364 | -0.029924 | 0.085181 | 22542 | 3.47E-01 | 5.45E-01 | 1.00E+00 |
| CC_Anterior | 0.026057 | 0.028953 | -0.030691 | 0.082805 | 22542 | 3.68E-01 | 5.52E-01 | 1.00E+00 |
| VesselVol_avg | 0.020237 | 0.024659 | -0.028093 | 0.068567 | 22542 | 4.12E-01 | 5.72E-01 | 1.00E+00 |
| PutamenVol_avg | -0.020722 | 0.025459 | -0.070621 | 0.029177 | 22542 | 4.16E-01 | 5.72E-01 | 1.00E+00 |
| FourthVent | 0.020254 | 0.030105 | -0.038751 | 0.079259 | 22542 | 5.01E-01 | 6.61E-01 | 1.00E+00 |
| CC_Mid_Posterior | -0.01674 | 0.030642 | -0.076798 | 0.043318 | 22542 | 5.85E-01 | 7.42E-01 | 1.00E+00 |
| CC_Posterior | -0.01239 | 0.030092 | -0.071369 | 0.046589 | 22542 | 6.81E-01 | 7.77E-01 | 1.00E+00 |
| PallidumVol_avg | -0.009758 | 0.023861 | -0.056524 | 0.037009 | 22542 | 6.83E-01 | 7.77E-01 | 1.00E+00 |
| ThirdVent | 0.010528 | 0.025268 | -0.038996 | 0.060052 | 22542 | 6.77E-01 | 7.77E-01 | 1.00E+00 |
| FifthVent | -0.012011 | 0.033798 | -0.078255 | 0.054232 | 22542 | 7.22E-01 | 7.95E-01 | 1.00E+00 |
| CaudateVol_avg | 0.007292 | 0.026896 | -0.045424 | 0.060007 | 22542 | 7.86E-01 | 8.37E-01 | 1.00E+00 |
| CerebralWhiteMatterVol_avg | -0.003202 | 0.016468 | -0.035478 | 0.029074 | 22542 | 8.46E-01 | 8.72E-01 | 1.00E+00 |
| non_WMhyperintensities | -0.005528 | 0.034203 | -0.072565 | 0.06151 | 22542 | 8.72E-01 | 8.72E-01 | 1.00E+00 |

***Supplementary Table 3:*** *Model summaries corresponding to the Supplementary Figure 2 forest plot. Model summaries include the brain regions of interest (coded ROIs), model estimates (Coef), standard errors, low and high confidence intervals, the sample size (N), the nominal p-value, and the p-values of both FDR and Bonferroni multiple comparisons test for each ROI.*

**<u>Supp. Figure 3:</u> The association between occupational pesticide exposure and regional brain volume differences (including fluid intelligence scores as a covariate)**

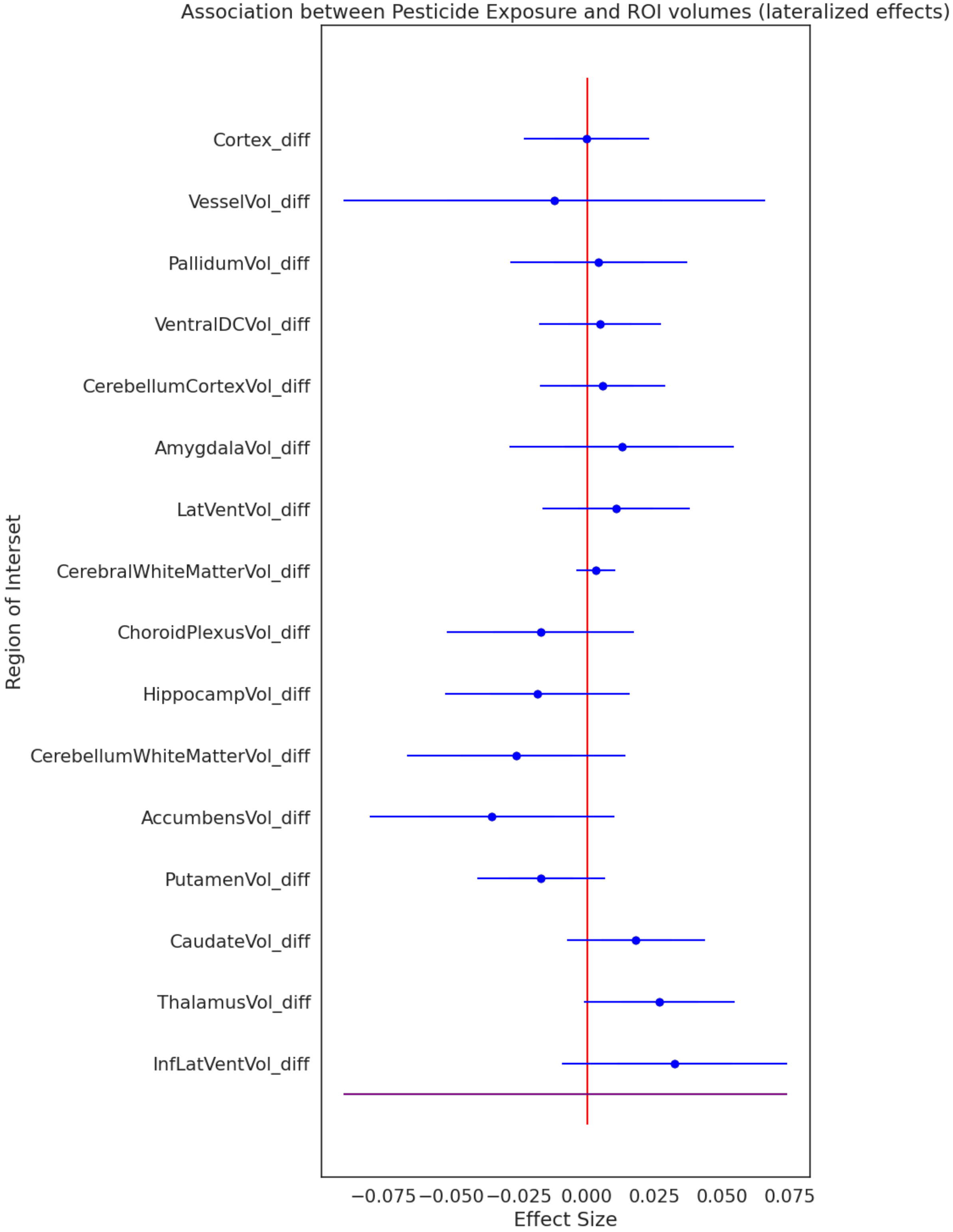

***Supplementary Figure 3:*** *Forest plot depicting the association between occupational pesticide exposure and ROI volume differences using generalized linear regression models. ROI volume differences were calculated by subtracting the right hemisphere ROI from the left hemisphere ROI. As such, a positive effect size corresponds to a right hemisphere lateralized effect and a negative effect size corresponds to a left hemisphere lateralized effect. Standard covariates were taken into account, including sex assigned at birth, age at the time of imaging, top 10 principal components of genetic ancestry (C1-C10), average fluid intelligence scores (across all visits), clinic site where imaging was conducted, and estimated total intracranial volume.*

#### Supplementary Table 4: Corresponding Model Summary for Supp. Figure 3

| Brain Region | Coef | Standard Errors | CI (low) | CI (high) | N | p_value | p_FDR | p_Bonferroni |
|---|---|---|---|---|---|---|---|---|
| InfLatVentVol_diff | 0.032433 | 0.021241 | -0.009199 | 0.074064 | 22542 | 1.27E-01 | 5.32E-01 | 1.00E+00 |
| ThalamusVol_diff | 0.02676 | 0.014189 | -0.001049 | 0.054569 | 22542 | 5.93E-02 | 5.32E-01 | 9.49E-01 |
| CaudateVol_diff | 0.018017 | 0.013014 | -0.007491 | 0.043525 | 22542 | 1.66E-01 | 5.32E-01 | 1.00E+00 |
| PutamenVol_diff | -0.016948 | 0.012043 | -0.040551 | 0.006655 | 22542 | 1.59E-01 | 5.32E-01 | 1.00E+00 |
| AccumbensVol_diff | -0.035137 | 0.023041 | -0.080296 | 0.010022 | 22542 | 1.27E-01 | 5.32E-01 | 1.00E+00 |
| CerebellumWhiteMatterVol_diff | -0.026121 | 0.020539 | -0.066376 | 0.014135 | 22542 | 2.03E-01 | 5.43E-01 | 1.00E+00 |
| HippocampVol_diff | -0.018375 | 0.017408 | -0.052493 | 0.015744 | 22542 | 2.91E-01 | 6.55E-01 | 1.00E+00 |
| ChoroidPlexusVol_diff | -0.017214 | 0.017581 | -0.051673 | 0.017244 | 22542 | 3.28E-01 | 6.55E-01 | 1.00E+00 |
| CerebralWhiteMatterVol_diff | 0.003202 | 0.003704 | -0.004057 | 0.010461 | 22542 | 3.87E-01 | 6.88E-01 | 1.00E+00 |
| LatVentVol_diff | 0.010671 | 0.013864 | -0.016502 | 0.037845 | 22542 | 4.41E-01 | 7.06E-01 | 1.00E+00 |
| AmygdalaVol_diff | 0.012807 | 0.021201 | -0.028747 | 0.054361 | 22542 | 5.46E-01 | 7.94E-01 | 1.00E+00 |
| CerebellumCortexVol_diff | 0.005646 | 0.011818 | -0.017516 | 0.028808 | 22542 | 6.33E-01 | 8.28E-01 | 1.00E+00 |
| VentralDCVol_diff | 0.004858 | 0.01151 | -0.017701 | 0.027416 | 22542 | 6.73E-01 | 8.28E-01 | 1.00E+00 |
| PallidumVol_diff | 0.004341 | 0.016634 | -0.028262 | 0.036943 | 22542 | 7.94E-01 | 8.47E-01 | 1.00E+00 |
| VesselVol_diff | -0.012056 | 0.039673 | -0.089813 | 0.065701 | 22542 | 7.61E-01 | 8.47E-01 | 1.00E+00 |
| Cortex_diff | -0.00019 | 0.011794 | -0.023307 | 0.022926 | 22542 | 9.87E-01 | 9.87E-01 | 1.00E+00 |

***Supplementary Table 4:*** *Model summaries corresponding to the Supplementary Figure 3 forest plot. Model summaries include the brain regions of interest (coded ROIs), model estimates (Coef), standard errors, low and high confidence intervals, the sample size (N), the nominal p-value, and the p-values of both FDR and Bonferroni multiple comparisons test for each ROI.*

**<u>Supp. Figure 4:</u> The association between occupational pesticide exposure and average regional brain volume differences (including all covariates)**

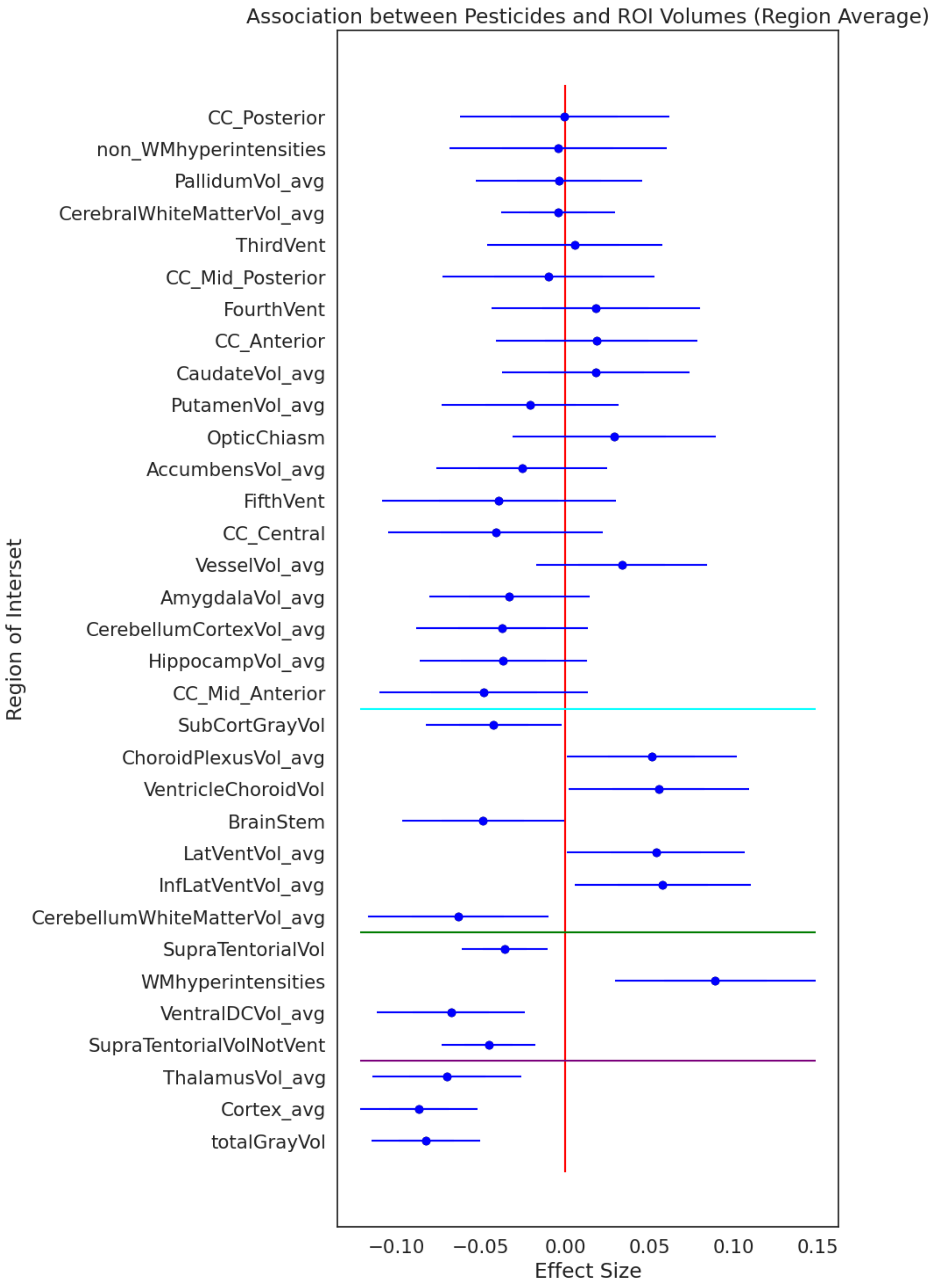

***Supplementary Figure 4****: Forest plot depicting the association between occupational pesticide exposure and ROI volume differences using generalized linear regression models. ROI volume values were averaged between hemispheres to account for bilateral effects. Standard covariates were taken into account, including sex assigned at birth, age at the time of imaging, top 10 principal components of genetic ancestry (C1-C10), average fluid intelligence scores (across all visits), Indices of Multiple Deprivation (health, education, and income), clinic site where imaging was conducted, and estimated total intracranial volume. The horizontal blue line represents nominal statistical significance, the horizontal green line represents the significance threshold for False Discovery Rate (FDR) correction, and the horizontal purple line represents the significance threshold for Bonferroni correction.*

#### Supplementary Table 5: Corresponding Model Summary for Supp. Figure 4

| Brain Region | Coef | Standard Errors | CI (low) | CI (high) | N | p_value | p_FDR | p_Bonferroni |
|---|---|---|---|---|---|---|---|---|
| totalGrayVol | -0.082515 | 0.016452 | -0.114759 | -0.050271 | 20515 | 5.29E-07 | 1.70E-05 | 1.70E-05 |
| Cortex_avg | -0.086693 | 0.017795 | -0.12157 | -0.051815 | 20515 | 1.11E-06 | 1.80E-05 | 3.70E-05 |
| ThalamusVol_avg | -0.069991 | 0.022559 | -0.114207 | -0.025776 | 20515 | 1.92E-03 | 1.58E-02 | 6.33E-02 |
| SupraTentorialVolNotVent | -0.045422 | 0.014275 | -0.073401 | -0.017444 | 20515 | 1.46E-03 | 1.58E-02 | 4.83E-02 |
| VentralDCVol_avg | -0.067775 | 0.022507 | -0.111889 | -0.023662 | 20515 | 2.60E-03 | 1.72E-02 | 8.59E-02 |
| WMhyperintensities | 0.08909 | 0.030277 | 0.029748 | 0.148432 | 20515 | 3.26E-03 | 1.79E-02 | 1.07E-01 |
| SupraTentorialVol | -0.035815 | 0.012932 | -0.061161 | -0.010469 | 20515 | 5.61E-03 | 2.65E-02 | 1.85E-01 |
| CerebellumWhiteMatterVol_avg | -0.063537 | 0.027308 | -0.11706 | -0.010014 | 20515 | 2.00E-02 | 8.24E-02 | 6.59E-01 |
| InfLatVentVol_avg | 0.057922 | 0.02662 | 0.005748 | 0.110095 | 20515 | 2.96E-02 | 1.08E-01 | 9.76E-01 |
| LatVentVol_avg | 0.053785 | 0.026841 | 0.001177 | 0.106392 | 20515 | 4.51E-02 | 1.11E-01 | 1.00E+00 |
| BrainStem | -0.048727 | 0.024527 | -0.096799 | -0.000655 | 20515 | 4.70E-02 | 1.11E-01 | 1.00E+00 |
| VentricleChoroidVol | 0.055577 | 0.027265 | 0.002139 | 0.109016 | 20515 | 4.15E-02 | 1.11E-01 | 1.00E+00 |
| ChoroidPlexusVol_avg | 0.051291 | 0.025768 | 0.000787 | 0.101796 | 20515 | 4.65E-02 | 1.11E-01 | 1.00E+00 |
| SubCortGrayVol | -0.0424 | 0.020464 | -0.082509 | -0.002291 | 20515 | 3.83E-02 | 1.11E-01 | 1.00E+00 |
| CC_Mid_Anterior | -0.048244 | 0.031527 | -0.110036 | 0.013548 | 20515 | 1.26E-01 | 2.77E-01 | 1.00E+00 |
| HippocampVol_avg | -0.036672 | 0.025358 | -0.086372 | 0.013028 | 20515 | 1.48E-01 | 2.88E-01 | 1.00E+00 |
| CerebellumCortexVol_avg | -0.037634 | 0.025965 | -0.088524 | 0.013255 | 20515 | 1.47E-01 | 2.88E-01 | 1.00E+00 |
| AmygdalaVol_avg | -0.033059 | 0.024176 | -0.080443 | 0.014325 | 20515 | 1.71E-01 | 3.14E-01 | 1.00E+00 |
| VesselVol_avg | 0.033524 | 0.025884 | -0.017209 | 0.084256 | 20515 | 1.95E-01 | 3.33E-01 | 1.00E+00 |
| CC_Central | -0.041316 | 0.032364 | -0.104749 | 0.022116 | 20515 | 2.02E-01 | 3.33E-01 | 1.00E+00 |
| FifthVent | -0.03933 | 0.035412 | -0.108736 | 0.030076 | 20515 | 2.67E-01 | 4.19E-01 | 1.00E+00 |

| | | | | | | | | |
|---|---|---|---|---|---|---|---|---|
| AccumbensVol_avg | -0.025731 | 0.025741 | -0.076183 | 0.024721 | 20515 | 3.18E-01 | 4.76E-01 | 1.00E+00 |
| OpticChiasm | 0.029114 | 0.03078 | -0.031215 | 0.089442 | 20515 | 3.44E-01 | 4.94E-01 | 1.00E+00 |
| PutamenVol_avg | -0.020674 | 0.026838 | -0.073275 | 0.031927 | 20515 | 4.41E-01 | 6.07E-01 | 1.00E+00 |
| CaudateVol_avg | 0.018408 | 0.028362 | -0.037181 | 0.073997 | 20515 | 5.16E-01 | 6.82E-01 | 1.00E+00 |
| CC_Anterior | 0.018764 | 0.030481 | -0.040978 | 0.078507 | 20515 | 5.38E-01 | 6.83E-01 | 1.00E+00 |
| FourthVent | 0.017944 | 0.03156 | -0.043912 | 0.079801 | 20515 | 5.70E-01 | 6.96E-01 | 1.00E+00 |
| CC_Mid_Posterior | -0.009975 | 0.032136 | -0.072961 | 0.053011 | 20515 | 7.56E-01 | 8.91E-01 | 1.00E+00 |
| ThirdVent | 0.005872 | 0.026498 | -0.046063 | 0.057806 | 20515 | 8.25E-01 | 9.07E-01 | 1.00E+00 |
| CerebralWhiteMatterVol_avg | -0.004224 | 0.017294 | -0.038121 | 0.029672 | 20515 | 8.07E-01 | 9.07E-01 | 1.00E+00 |
| PallidumVol_avg | -0.003605 | 0.02513 | -0.052858 | 0.045648 | 20515 | 8.86E-01 | 9.23E-01 | 1.00E+00 |
| non_WMhyperintensities | -0.004338 | 0.03287 | -0.068762 | 0.060086 | 20515 | 8.95E-01 | 9.23E-01 | 1.00E+00 |
| CC_Posterior | -0.000502 | 0.03168 | -0.062594 | 0.06159 | 20515 | 9.87E-01 | 9.87E-01 | 1.00E+00 |

***Supplementary Table 5:*** *Model summaries corresponding to the Supplementary Figure 4 forest plot. Model summaries include the brain regions of interest (coded ROIs), model estimates (Coef), standard errors, low and high confidence intervals, the sample size (N), the nominal p-value, and the p-values of both FDR and Bonferroni multiple comparisons test for each ROI.*

**<u>Supplementary Figure 5:</u> The association between occupational pesticide exposure and regional brain volume differences (including all covariates)**

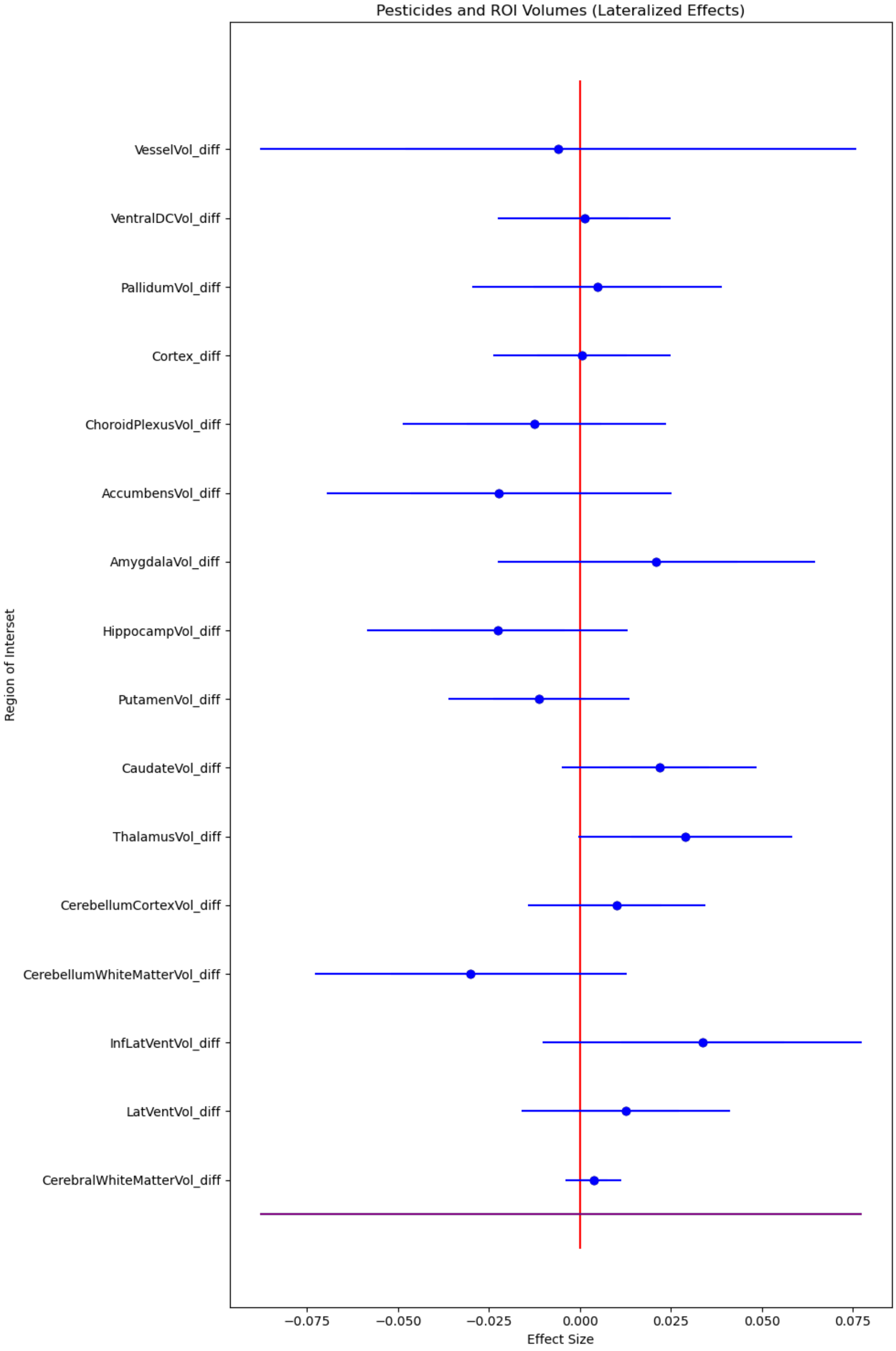

***Supplementary Figure 5****: Forest plot depicting the association between occupational pesticide exposure and ROI volume differences using generalized linear regression models. ROI volume differences were calculated by subtracting the right hemisphere ROI from the left hemisphere ROI. As such, a positive effect size corresponds to a right hemisphere lateralized effect and a negative effect size corresponds to a left hemisphere lateralized effect. Standard covariates were taken into account including sex assigned at birth, age at the time of imaging, top 10 principal components of genetic ancestry (C1-C10), average fluid intelligence scores (across all visits), Indices of Multiple Deprivation (health, education, and income), clinic site where imaging was conducted, and estimated total intracranial volume.*

#### Supplementary Table 6: Corresponding Model Summary for Supp. Figure 5

| Brain Region | Coef | Standard Errors | CI (low) | CI (high) | N | p_value | p_FDR | p_Bonferroni |
|---|---|---|---|---|---|---|---|---|
| CerebralWhiteMatterVol_diff | 0.003719 | 0.003881 | -0.003888 | 0.011326 | 20515 | 3.38E-01 | 6.11E-01 | 1.00E+00 |
| LatVentVol_diff | 0.012618 | 0.014571 | -0.01594 | 0.041176 | 20515 | 3.86E-01 | 6.11E-01 | 1.00E+00 |
| InfLatVentVol_diff | 0.033631 | 0.022366 | -0.010205 | 0.077467 | 20515 | 1.33E-01 | 6.11E-01 | 1.00E+00 |
| CerebellumWhiteMatterVol_diff | -0.030067 | 0.021839 | -0.07287 | 0.012737 | 20515 | 1.69E-01 | 6.11E-01 | 1.00E+00 |
| CerebellumCortexVol_diff | 0.010034 | 0.012449 | -0.014365 | 0.034433 | 20515 | 4.20E-01 | 6.11E-01 | 1.00E+00 |
| ThalamusVol_diff | 0.028947 | 0.01496 | -0.000375 | 0.058269 | 20515 | 5.30E-02 | 6.11E-01 | 8.48E-01 |
| CaudateVol_diff | 0.021787 | 0.013636 | -0.00494 | 0.048514 | 20515 | 1.10E-01 | 6.11E-01 | 1.00E+00 |
| PutamenVol_diff | -0.011266 | 0.01269 | -0.036139 | 0.013607 | 20515 | 3.75E-01 | 6.11E-01 | 1.00E+00 |
| HippocampVol_diff | -0.022594 | 0.018273 | -0.05841 | 0.013221 | 20515 | 2.16E-01 | 6.11E-01 | 1.00E+00 |
| AmygdalaVol_diff | 0.020994 | 0.02226 | -0.022635 | 0.064623 | 20515 | 3.46E-01 | 6.11E-01 | 1.00E+00 |
| AccumbensVol_diff | -0.022246 | 0.024169 | -0.069617 | 0.025124 | 20515 | 3.57E-01 | 6.11E-01 | 1.00E+00 |
| ChoroidPlexusVol_diff | -0.012572 | 0.018511 | -0.048852 | 0.023708 | 20515 | 4.97E-01 | 6.63E-01 | 1.00E+00 |
| Cortex_diff | 0.000538 | 0.012384 | -0.023733 | 0.02481 | 20515 | 9.65E-01 | 9.65E-01 | 1.00E+00 |
| PallidumVol_diff | 0.004735 | 0.017514 | -0.029593 | 0.039062 | 20515 | 7.87E-01 | 9.65E-01 | 1.00E+00 |
| VentralDCVol_diff | 0.001184 | 0.012094 | -0.02252 | 0.024887 | 20515 | 9.22E-01 | 9.65E-01 | 1.00E+00 |
| VesselVol_diff | -0.006108 | 0.041804 | -0.088043 | 0.075827 | 20515 | 8.84E-01 | 9.65E-01 | 1.00E+00 |

***Supplementary Table 6:*** *Model summaries corresponding to the Supplementary Figure 5 forest plot. Model summaries include the brain regions of interest (coded ROIs), model estimates (Coef), standard errors, low and high confidence intervals, the sample size (N), the nominal p-value, and the p-values of both FDR and Bonferroni multiple comparisons test for each ROI.*

**<u>Supplementary Figure 6:</u> The effect of fluid intelligence on the relationship between occupational pesticide exposure and ROI volumes (regional average and difference)**

**A**

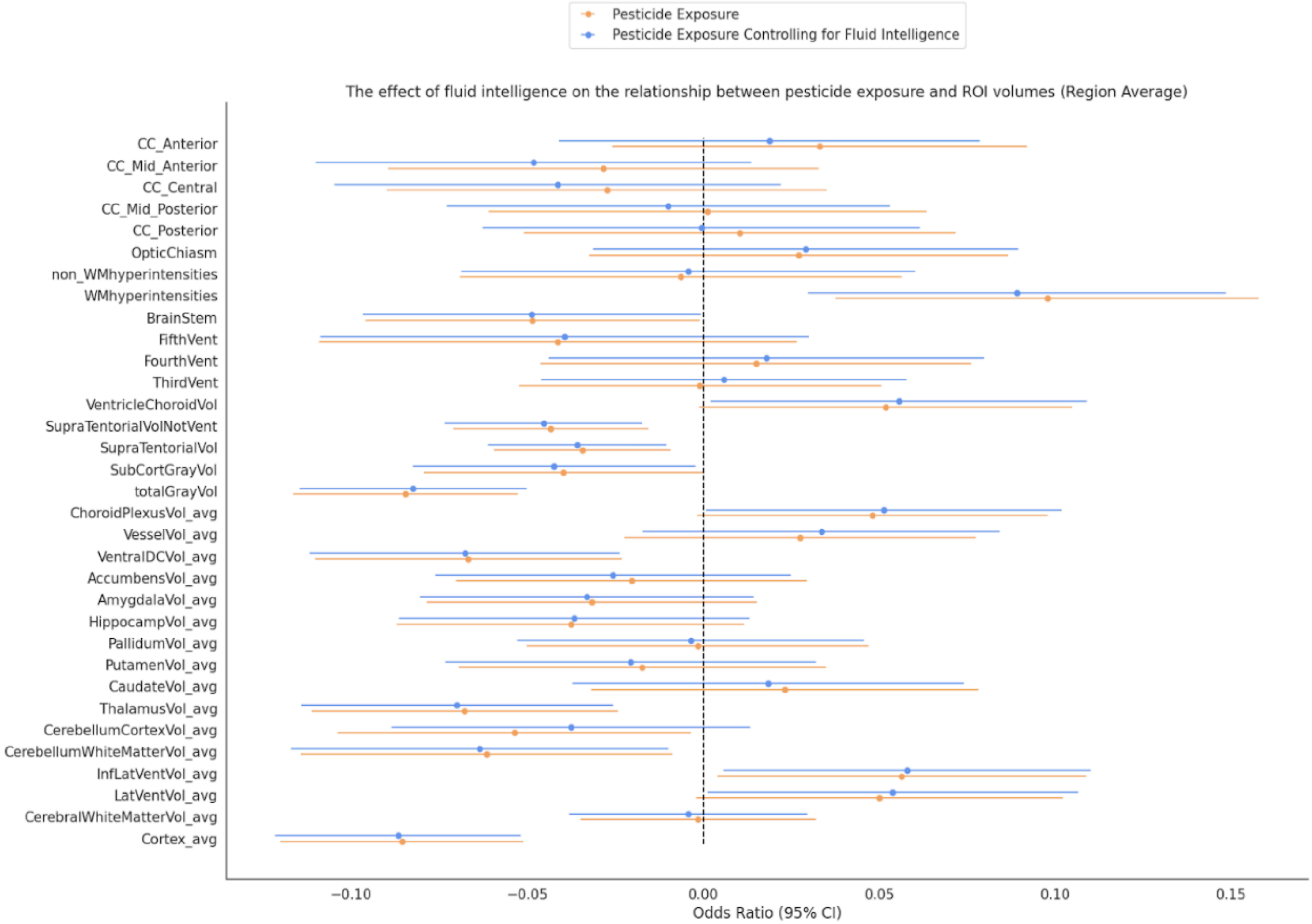

**B**

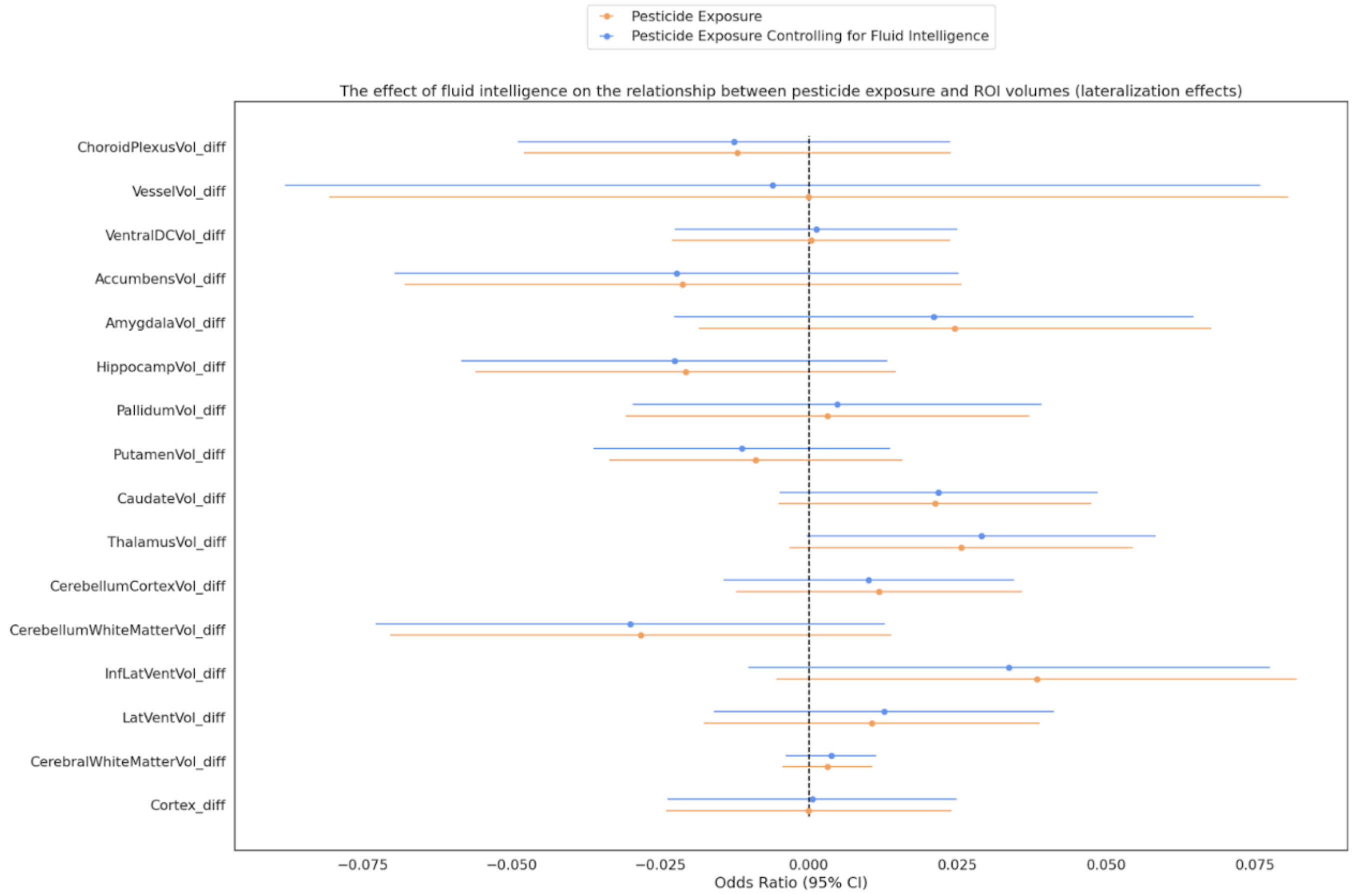
Pesticide Exposure
Pesticide Exposure Controlling for Fluid Intelligence
The effect of fluid intelligence on the relationship between pesticide exposure and ROI volumes (lateralization effects)
ChoroidPlexusVol_diff
VesselVol_diff
VentralDCVol_diff
AccumbensVol_diff
AmygdalaVol_diff
HippocampVol_diff
PallidumVol_diff
PutamenVol_diff
CaudateVol_diff
ThalamusVol_diff
CerebellumCortexVol_diff
CerebellumWhiteMatterVol_diff
InfLatVentVol_diff
LatVentVol_diff
CerebralWhiteMatterVol_diff
Cortex_diff
−0.075
−0.050
−0.025
0.000
0.025
0.050
0.075
Odds Ratio (95% CI)

**C**

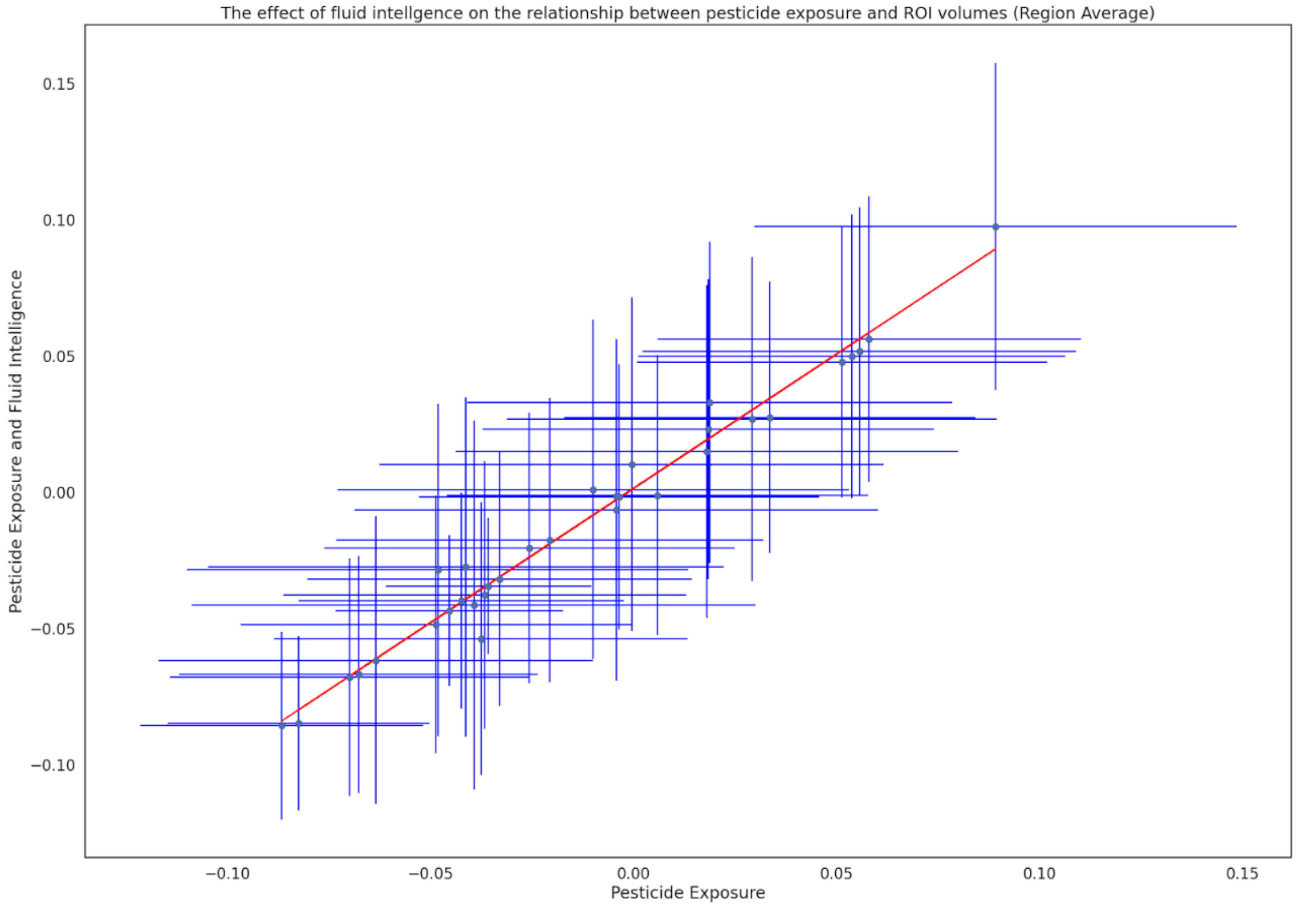


***Supplementary Figure 6****: Forest plots depicting the effect of fluid intelligence scores as a covariate on the association between occupational pesticide exposure and ROI volumetric differences. Model estimates and confidence intervals highlighted in orange account for the following covariates: age at time of imaging, sex assigned at birth, top 10 principal components of genetic ancestry (C1-C10),*

***A:*** *Condensed forest plot depicting the effect of fluid intelligence scores on the association between occupational pesticide exposure and average ROI volume values. Model estimates and confidence intervals highlighted in blue depict the relationship between pesticide exposure and ROI average volume with fluid intelligence scores as a covariate, whereas those in orange depict the relationship between pesticide exposure and ROI average volumes without taking into account fluid intelligence scores. We observe no significant differences in model estimates.*

***B:*** *Condensed forest plot depicting the effect of fluid intelligence scores on the association between occupational pesticide exposure and lateralized ROI volume values (values* *are the difference between regional hemispheres). Model estimates and confidence intervals highlighted in blue depict the relationship between pesticide exposure and ROI volume lateralization values with fluid intelligence scores as a covariate, whereas those in orange depict the relationship between pesticide exposure and ROI volume lateralization values without taking into account fluid intelligence scores. We observe no significant differences in model estimates.*

***C:*** *Scatterplot depicting the effect of fluid intelligence on the relationship between occupational pesticide exposure and average ROI volume values (Forest Plot A in Supplementary Figure 6). The x-axis depicts the association between pesticide exposure and average ROI volume values, and the y-axis depicts the association between pesticide exposure and average ROI volume values with fluid intelligence scores as a covariate. The blue dots on the scatterplot depict model estimates, and the blue horizontal and vertical lines represent confidence intervals for the respective general linear model. The red line depicts the line of best fit.*

**Supplementary Table 7: Corresponding Model Summary for Supplementary Figures 6A and 6C - Pesticide Exposure (Region Average)**

| Brain Region | Coef | Standard Errors | CI (low) | CI (high) | N | p_value | p_FDR | p_Bonferroni |
|---|---|---|---|---|---|---|---|---|
| totalGrayVol | -0.084702 | 0.016276 | -0.116602 | -0.052803 | 21049 | 1.95E-07 | 6.00E-06 | 6.00E-06 |
| Cortex_avg | -0.085617 | 0.017594 | -0.120099 | -0.051134 | 21049 | 1.14E-06 | 1.90E-05 | 3.80E-05 |
| ThalamusVol_avg | -0.067792 | 0.022245 | -0.111391 | -0.024194 | 21049 | 2.31E-03 | 1.47E-02 | 7.61E-02 |
| WMhyperintensities | 0.097721 | 0.030652 | 0.037645 | 0.157798 | 21049 | 1.43E-03 | 1.47E-02 | 4.73E-02 |
| SupraTentorialVolNotVent | -0.04331 | 0.014121 | -0.070986 | -0.015634 | 21049 | 2.16E-03 | 1.47E-02 | 7.13E-02 |
| VentralDCVol_avg | -0.066755 | 0.022223 | -0.110311 | -0.023199 | 21049 | 2.67E-03 | 1.47E-02 | 8.80E-02 |
| SupraTentorialVol | -0.034321 | 0.012779 | -0.059368 | -0.009274 | 21049 | 7.24E-03 | 3.41E-02 | 2.39E-01 |
| CerebellumWhiteMatterVol_avg | -0.061579 | 0.02697 | -0.114439 | -0.00872 | 21049 | 2.24E-02 | 9.25E-02 | 7.40E-01 |
| InfLatVentVol_avg | 0.056401 | 0.026754 | 0.003964 | 0.108839 | 21049 | 3.50E-02 | 1.20E-01 | 1.00E+00 |
| CerebellumCortexVol_avg | -0.053652 | 0.02562 | -0.103867 | -0.003437 | 21049 | 3.62E-02 | 1.20E-01 | 1.00E+00 |
| LatVentVol_avg | 0.049989 | 0.026579 | -0.002105 | 0.102084 | 21049 | 6.00E-02 | 1.32E-01 | 1.00E+00 |
| BrainStem | -0.048509 | 0.024197 | -0.095934 | -0.001083 | 21049 | 4.50E-02 | 1.32E-01 | 1.00E+00 |
| VentricleChoroidVol | 0.051812 | 0.027007 | -0.00112 | 0.104744 | 21049 | 5.50E-02 | 1.32E-01 | 1.00E+00 |
| ChoroidPlexusVol_avg | 0.047962 | 0.025387 | -0.001797 | 0.09772 | 21049 | 5.89E-02 | 1.32E-01 | 1.00E+00 |
| SubCortGrayVol | -0.039721 | 0.020257 | -0.079424 | -0.000019 | 21049 | 4.99E-02 | 1.32E-01 | 1.00E+00 |
| HippocampVol_avg | -0.03768 | 0.025119 | -0.086911 | 0.011552 | 21049 | 1.34E-01 | 2.76E-01 | 1.00E+00 |
| AmygdalaVol_avg | -0.031675 | 0.02388 | -0.078479 | 0.015128 | 21049 | 1.85E-01 | 3.59E-01 | 1.00E+00 |
| FifthVent | -0.041289 | 0.034593 | -0.10909 | 0.026511 | 21049 | 2.33E-01 | 4.27E-01 | 1.00E+00 |

| | | | | | | | | |
|---|---|---|---|---|---|---|---|---|
| VesselVol_avg | 0.027552 | 0.02547 | -0.022369 | 0.077473 | 21049 | 2.79E-01 | 0.46097 | 1 |
| CC_Anterior | 0.033091 | 0.030072 | -0.025848 | 0.092031 | 21049 | 2.71E-01 | 0.46097 | 1 |
| CC_Mid_Anterior | -0.028419 | 0.03114 | -0.089453 | 0.032615 | 21049 | 3.61E-01 | 0.558356 | 1 |
| CaudateVol_avg | 0.023153 | 0.028086 | -0.031895 | 0.078201 | 21049 | 4.10E-01 | 0.558356 | 1 |
| OpticChiasm | 0.027104 | 0.030383 | -0.032446 | 0.086653 | 21049 | 3.72E-01 | 0.558356 | 1 |
| CC_Central | -0.027389 | 0.03185 | -0.089814 | 0.035035 | 21049 | 3.90E-01 | 0.558356 | 1 |
| AccumbensVol_avg | -0.020355 | 0.025405 | -0.070148 | 0.029438 | 21049 | 4.23E-01 | 0.558356 | 1 |
| PutamenVol_avg | -0.017348 | 0.026604 | -0.069492 | 0.034795 | 21049 | 5.14E-01 | 0.652815 | 1 |
| FourthVent | 0.014975 | 0.03119 | -0.046156 | 0.076107 | 21049 | 6.31E-01 | 0.77139 | 1 |
| CC_Posterior | 0.010369 | 0.031284 | -0.050947 | 0.071684 | 21049 | 7.40E-01 | 0.872516 | 1 |
| non_WMhyperintensities | -0.006368 | 0.032012 | -0.069111 | 0.056375 | 21049 | 8.42E-01 | 0.958504 | 1 |
| PallidumVol_avg | -0.001613 | 0.024817 | -0.050252 | 0.047027 | 21049 | 9.48E-01 | 0.971397 | 1 |
| ThirdVent | -0.000941 | 0.026256 | -0.052403 | 0.05052 | 21049 | 9.71E-01 | 0.971397 | 1 |
| CC_Mid_Posterior | 0.001163 | 0.031761 | -0.061087 | 0.063414 | 21049 | 9.71E-01 | 0.971397 | 1 |
| CerebralWhiteMatterVol_avg | -0.001456 | 0.017065 | -0.034904 | 0.031991 | 21049 | 9.32E-01 | 0.971397 | 1 |

***Supplementary Table 7:*** *Model summaries corresponding to the Supplementary Figure 6A and 6C forest plots. Model summaries include the brain regions of interest (coded ROIs), model estimates (Coef), standard errors, low and high confidence intervals, the sample size (N), the nominal p-value, and the p-values of both FDR and Bonferroni multiple comparisons test for each ROI.*

#### Supplementary Table 8: Corresponding Model Summary for Figures 6A and 6C: Pesticide Exposure Controlling for Fluid Intelligence (Region Average)

| Brain Region | Coef | Standard Errors | CI (low) | CI (high) | N | p_value | p_FDR | p_Bonferroni |
|---|---|---|---|---|---|---|---|---|
| totalGrayVol | -0.082515 | 0.016452 | -0.114759 | -0.050271 | 20515 | 5.29E-07 | 1.70E-05 | 1.70E-05 |
| Cortex_avg | -0.086693 | 0.017795 | -0.12157 | -0.051815 | 20515 | 1.11E-06 | 1.80E-05 | 3.70E-05 |
| ThalamusVol_avg | -0.069991 | 0.022559 | -0.114207 | -0.025776 | 20515 | 1.92E-03 | 1.58E-02 | 6.33E-02 |
| SupraTentorialVolNotVent | -0.045422 | 0.014275 | -0.073401 | -0.017444 | 20515 | 1.46E-03 | 1.58E-02 | 4.83E-02 |
| VentralDCVol_avg | -0.067775 | 0.022507 | -0.111889 | -0.023662 | 20515 | 2.60E-03 | 1.72E-02 | 8.59E-02 |
| WMhyperintensities | 0.08909 | 0.030277 | 0.029748 | 0.148432 | 20515 | 3.26E-03 | 1.79E-02 | 1.07E-01 |
| SupraTentorialVol | -0.035815 | 0.012932 | -0.061161 | -0.010469 | 20515 | 5.61E-03 | 2.65E-02 | 1.85E-01 |
| CerebellumWhiteMatterVol_avg | -0.063537 | 0.027308 | -0.11706 | -0.010014 | 20515 | 2.00E-02 | 8.24E-02 | 6.59E-01 |
| InfLatVentVol_avg | 0.057922 | 0.02662 | 0.005748 | 0.110095 | 20515 | 2.96E-02 | 1.08E-01 | 9.76E-01 |
| LatVentVol_avg | 0.053785 | 0.026841 | 0.001177 | 0.106392 | 20515 | 4.51E-02 | 1.11E-01 | 1.00E+00 |

| | | | | | | | | |
|---|---|---|---|---|---|---|---|---|
| BrainStem | -0.048727 | 0.024527 | -0.096799 | -0.000655 | 20515 | 4.70E-02 | 1.11E-01 | 1.00E+00 |
| VentricleChoroidVol | 0.055577 | 0.027265 | 0.002139 | 0.109016 | 20515 | 4.15E-02 | 1.11E-01 | 1.00E+00 |
| ChoroidPlexusVol_avg | 0.051291 | 0.025768 | 0.000787 | 0.101796 | 20515 | 4.65E-02 | 1.11E-01 | 1.00E+00 |
| SubCortGrayVol | -0.0424 | 0.020464 | -0.082509 | -0.002291 | 20515 | 3.83E-02 | 1.11E-01 | 1.00E+00 |
| CC_Mid_Anterior | -0.048244 | 0.031527 | -0.110036 | 0.013548 | 20515 | 1.26E-01 | 2.77E-01 | 1.00E+00 |
| HippocampVol_avg | -0.036672 | 0.025358 | -0.086372 | 0.013028 | 20515 | 1.48E-01 | 2.88E-01 | 1.00E+00 |
| CerebellumCortexVol_avg | -0.037634 | 0.025965 | -0.088524 | 0.013255 | 20515 | 1.47E-01 | 2.88E-01 | 1.00E+00 |
| AmygdalaVol_avg | -0.033059 | 0.024176 | -0.080443 | 0.014325 | 20515 | 1.71E-01 | 3.14E-01 | 1.00E+00 |
| VesselVol_avg | 0.033524 | 0.025884 | -0.017209 | 0.084256 | 20515 | 1.95E-01 | 3.33E-01 | 1.00E+00 |
| CC_Central | -0.041316 | 0.032364 | -0.104749 | 0.022116 | 20515 | 2.02E-01 | 3.33E-01 | 1.00E+00 |
| FifthVent | -0.03933 | 0.035412 | -0.108736 | 0.030076 | 20515 | 2.67E-01 | 4.19E-01 | 1.00E+00 |
| AccumbensVol_avg | -0.025731 | 0.025741 | -0.076183 | 0.024721 | 20515 | 3.18E-01 | 4.76E-01 | 1.00E+00 |
| OpticChiasm | 0.029114 | 0.03078 | -0.031215 | 0.089442 | 20515 | 3.44E-01 | 4.94E-01 | 1.00E+00 |
| PutamenVol_avg | -0.020674 | 0.026838 | -0.073275 | 0.031927 | 20515 | 4.41E-01 | 6.07E-01 | 1.00E+00 |
| CaudateVol_avg | 0.018408 | 0.028362 | -0.037181 | 0.073997 | 20515 | 5.16E-01 | 6.82E-01 | 1.00E+00 |
| CC_Anterior | 0.018764 | 0.030481 | -0.040978 | 0.078507 | 20515 | 5.38E-01 | 6.83E-01 | 1.00E+00 |
| FourthVent | 0.017944 | 0.03156 | -0.043912 | 0.079801 | 20515 | 5.70E-01 | 6.96E-01 | 1.00E+00 |
| CC_Mid_Posterior | -0.009975 | 0.032136 | -0.072961 | 0.053011 | 20515 | 7.56E-01 | 8.91E-01 | 1.00E+00 |
| ThirdVent | 0.005872 | 0.026498 | -0.046063 | 0.057806 | 20515 | 8.25E-01 | 9.07E-01 | 1.00E+00 |
| CerebralWhiteMatterVol_avg | -0.004224 | 0.017294 | -0.038121 | 0.029672 | 20515 | 8.07E-01 | 9.07E-01 | 1.00E+00 |
| PallidumVol_avg | -0.003605 | 0.02513 | -0.052858 | 0.045648 | 20515 | 8.86E-01 | 9.23E-01 | 1.00E+00 |
| non_WMhyperintensities | -0.004338 | 0.03287 | -0.068762 | 0.060086 | 20515 | 8.95E-01 | 9.23E-01 | 1.00E+00 |
| CC_Posterior | -0.000502 | 0.03168 | -0.062594 | 0.06159 | 20515 | 9.87E-01 | 9.87E-01 | 1.00E+00 |

***Supplementary Table 8:*** *Model summaries corresponding to the Supplementary Figure 6A and 6C forest plots. Model summaries include the brain regions of interest (coded ROIs), model estimates (Coef), standard errors, low and high confidence intervals, the sample size (N), the nominal p-value, and the p-values of both FDR and Bonferroni multiple comparisons test for each ROI.*

#### Supplementary Table 9: Corresponding Model Summary for Figure 6B: Pesticide Exposure (Lateralization Effects)

| Brain Region | Estimates | Standard Errors | CI (low) | CI (high) | N | p_value | p_FDR | p_Bonferroni |
|---|---|---|---|---|---|---|---|---|
| InfLatVentVol_diff | 0.038265 | 0.022315 | -0.005472 | 0.082002 | 21049 | 8.64E-02 | 6.13E-01 | 1.00E+00 |
| ThalamusVol_diff | 0.025591 | 0.014727 | -0.003273 | 0.054454 | 21049 | 8.23E-02 | 6.13E-01 | 1.00E+00 |

| | | | | | | | | |
|---|---|---|---|---|---|---|---|---|
| CaudateVol_diff | 0.021163 | 0.013424 | -0.005148 | 0.047474 | 21049 | 1.15E-01 | 6.13E-01 | 1.00E+00 |
| CerebralWhiteMatterVol_diff | 0.003075 | 0.003852 | -0.004474 | 0.010624 | 21049 | 4.25E-01 | 6.80E-01 | 1.00E+00 |
| LatVentVol_diff | 0.010582 | 0.014373 | -0.017589 | 0.038753 | 21049 | 4.62E-01 | 6.80E-01 | 1.00E+00 |
| CerebellumWhiteMatterVol_diff | -0.028248 | 0.021484 | -0.070356 | 0.013861 | 21049 | 1.89E-01 | 6.80E-01 | 1.00E+00 |
| CerebellumCortexVol_diff | 0.011743 | 0.012256 | -0.012279 | 0.035765 | 21049 | 3.38E-01 | 6.80E-01 | 1.00E+00 |
| PutamenVol_diff | -0.008924 | 0.01257 | -0.033561 | 0.015712 | 21049 | 4.78E-01 | 6.80E-01 | 1.00E+00 |
| HippocampVol_diff | -0.020739 | 0.018017 | -0.056052 | 0.014574 | 21049 | 2.50E-01 | 6.80E-01 | 1.00E+00 |
| AmygdalaVol_diff | 0.024501 | 0.021973 | -0.018565 | 0.067567 | 21049 | 2.65E-01 | 6.80E-01 | 1.00E+00 |
| AccumbensVol_diff | -0.021238 | 0.023867 | -0.068016 | 0.025539 | 21049 | 3.74E-01 | 6.80E-01 | 1.00E+00 |
| ChoroidPlexusVol_diff | -0.012037 | 0.018285 | -0.047875 | 0.023801 | 21049 | 5.10E-01 | 6.80E-01 | 1.00E+00 |
| Cortex_diff | -0.000037 | 0.012223 | -0.023993 | 0.023919 | 21049 | 9.98E-01 | 9.99E-01 | 1.00E+00 |
| PallidumVol_diff | 0.003087 | 0.017298 | -0.030815 | 0.03699 | 21049 | 8.58E-01 | 9.99E-01 | 1.00E+00 |
| VentralDCVol_diff | 0.000342 | 0.011929 | -0.023039 | 0.023722 | 21049 | 9.77E-01 | 9.99E-01 | 1.00E+00 |
| VesselVol_diff | -0.000036 | 0.041105 | -0.080599 | 0.080528 | 21049 | 9.99E-01 | 9.99E-01 | 1.00E+00 |

***Supplementary Table 9:*** *Model summaries corresponding to the Supplementary Figure 6B forest plot. Model summaries include the brain regions of interest (coded ROIs), model estimates (Coef), standard errors, low and high confidence intervals, the sample size (N), the nominal p-value, and the p-values of both FDR and Bonferroni multiple comparisons test for each ROI.*

#### Supplementary Table 10: Corresponding Model Summary for Figure B: Pesticide Exposure Controlling for Fluid Intelligence (Lateralization Effects)

| Brain Region | Coef | Standard Errors | N | CI (low) | CI (high) | p_value | p_FDR | p_Bonferroni |
|---|---|---|---|---|---|---|---|---|
| CerebralWhiteMatterVol_diff | 0.003719 | 0.003881 | 20515 | -0.003888 | 0.011326 | 3.38E-01 | 6.11E-01 | 1.00E+00 |
| LatVentVol_diff | 0.012618 | 0.014571 | 20515 | -0.01594 | 0.041176 | 3.86E-01 | 6.11E-01 | 1.00E+00 |
| InfLatVentVol_diff | 0.033631 | 0.022366 | 20515 | -0.010205 | 0.077467 | 1.33E-01 | 6.11E-01 | 1.00E+00 |
| CerebellumWhiteMatterVol_diff | -0.030067 | 0.021839 | 20515 | -0.07287 | 0.012737 | 1.69E-01 | 6.11E-01 | 1.00E+00 |
| CerebellumCortexVol_diff | 0.010034 | 0.012449 | 20515 | -0.014365 | 0.034433 | 4.20E-01 | 6.11E-01 | 1.00E+00 |
| ThalamusVol_diff | 0.028947 | 0.01496 | 20515 | -0.000375 | 0.058269 | 5.30E-02 | 6.11E-01 | 8.48E-01 |
| CaudateVol_diff | 0.021787 | 0.013636 | 20515 | -0.00494 | 0.048514 | 1.10E-01 | 6.11E-01 | 1.00E+00 |
| PutamenVol_diff | -0.011266 | 0.01269 | 20515 | -0.036139 | 0.013607 | 3.75E-01 | 6.11E-01 | 1.00E+00 |
| HippocampVol_diff | -0.022594 | 0.018273 | 20515 | -0.05841 | 0.013221 | 2.16E-01 | 6.11E-01 | 1.00E+00 |
| AmygdalaVol_diff | 0.020994 | 0.02226 | 20515 | -0.022635 | 0.064623 | 3.46E-01 | 6.11E-01 | 1.00E+00 |
| AccumbensVol_diff | -0.022246 | 0.024169 | 20515 | -0.069617 | 0.025124 | 3.57E-01 | 6.11E-01 | 1.00E+00 |
| ChoroidPlexusVol_diff | -0.012572 | 0.018511 | 20515 | -0.048852 | 0.023708 | 4.97E-01 | 6.63E-01 | 1.00E+00 |

| | | | | | | | | |
|---|---|---|---|---|---|---|---|---|
| Cortex_diff | 0.000538 | 0.012384 | 20515 | -0.023733 | 0.02481 | 9.65E-01 | 9.65E-01 | 1.00E+00 |
| PallidumVol_diff | 0.004735 | 0.017514 | 20515 | -0.029593 | 0.039062 | 7.87E-01 | 9.65E-01 | 1.00E+00 |
| VentralDCVol_diff | 0.001184 | 0.012094 | 20515 | -0.02252 | 0.024887 | 9.22E-01 | 9.65E-01 | 1.00E+00 |
| VesselVol_diff | -0.006108 | 0.041804 | 20515 | -0.088043 | 0.075827 | 8.84E-01 | 9.65E-01 | 1.00E+00 |

***Supplementary Table 10:*** *Model summaries corresponding to the Supplementary Figure 6B forest plot. Model summaries include the brain regions of interest (coded ROIs), model estimates (Coef), standard errors, low and high confidence intervals, the sample size (N), the nominal p-value, and the p-values of both FDR and Bonferroni multiple comparisons test for each ROI.*
